# Explainable Multimodal AI for In-Hospital Detection of Lung Cancer and Heart Failure Using Routine Data: Development, Internal Validation, and Simulated Federated Evaluation

**DOI:** 10.64898/2026.10.22.26366034

**Authors:** Mostak Ahmed, Md Mostafizur Rahman, Sabera Tabassum Wahid, Golam Rabbi, Al Rafy, Golam Sarwar Khondoker, Shovit Dutta, Mohammad Amir Saadat, Md. Masuqul Haque

## Abstract

**Objective:** To determine whether combining four routine data sources improves in-hospital detection of lung cancer and heart failure, which sources matter, whether explanations are faithful, whether models are fair, and how much benefit federated training retains.

**Methods:** This retrospective single-centre study linked MIMIC-IV admissions (73,637 patients) to chest radiographs, radiology reports, electrocardiograms (ECGs) and laboratory tests from the first 48 hours. Excluding prior cases, we compared single-source models, gradient boosting and early, late and attention-based fusion, tested explanations for faithfulness and weight dependence, audited subgroup performance, and simulated five hospitals for federated training.

**Results:** Lung cancer was coded in 1.7% and heart failure in 15.6%. Attention fusion reached an area under the receiver operating characteristic curve (AUROC) of 0.833 (95% CI 0.809–0.859) for lung cancer and 0.885 (0.878–0.892) for heart failure, against 0.753 and 0.794 for gradient boosting on laboratory data (both p < 0.001); early and late fusion performed similarly. Lung cancer relied most on laboratory data and reports, heart failure on the ECG. Explanations passed a deletion test; weight randomisation of absolute attributions passed for ECG but not laboratory explanations. Heart-failure discrimination fell with age. Federated training recovered 95% of the centralised gain over local training for heart failure; for lung cancer, federated averaging (FedAvg) recovered 51% and FedProx 82%.

**Conclusion:** Fusing routine data improved in-hospital detection of both diseases over laboratory data alone, and which data matter depends on the disease. Explanations should be checked per source, and performance in older patients monitored. External validation is needed.

## 1. Introduction

Chronic diseases are often recognised late, even when the evidence for them is already in the record. Lung cancer is the most commonly diagnosed cancer and the leading cause of cancer death worldwide, with about 2.5 million new cases and 1.8 million deaths in 2022 [1]. A large share of cases is first diagnosed after an emergency presentation, with poor survival [2]. Lung cancers are also missed on chest radiographs taken for other reasons [3]. Heart failure affects more than 60 million people and is a leading cause of hospital admission [4]; when it is coded at discharge, its signs are often present early in the stay. For both diseases the early evidence (a nodule or mass on the first radiograph, a phrase in the radiologist’s report, an abnormal electrocardiogram (ECG), anaemia or hyponatraemia) is collected within hours of arrival. But it sits in separate systems and is rarely read together.

Combining these sources could flag both diseases before discharge. Multimodal models are usually judged on discrimination alone, while which sources matter, whether explanations reflect the model, whether performance is equal across groups and what federated training costs are evaluated separately, if at all (Section 2). We therefore built a reusable, leakage-controlled evaluation framework that addresses these questions in one design, and applied it to two chronic diseases with different patho-physiology, lung cancer as the primary outcome and heart failure as the secondary outcome, using data recorded in the first 48 hours after hospital arrival. By in-hospital detection we mean flagging, from these data and before discharge, disease that is coded at discharge. Our contributions are:

1. A leakage-controlled cohort that links Medical Information Mart for Intensive Care (MIMIC)-IV laboratory results, chest radiographs, radiology reports and ECGs by patient and time, excludes patients already known to have the disease, and is checked for leakage automatically.
2. A comparison of single-source and fused models for both diseases, with ablations that measure what each source adds and a pre-specified primary hypothesis.
3. Explanations that are tested for faithfulness and for dependence on the learned weights, not merely displayed.
4. A fairness audit across sex, age, race and ethnicity, and insurance.
5. A federated evaluation across simulated hospitals with unequal data.

Reporting follows the TRIPOD+AI guideline (Transparent Reporting of a multivariable prediction model for Individual Prognosis Or Diagnosis, updated for artificial intelligence) [5], and the completed checklist is provided as a separate supplementary file.

## 2. Related work

### 2.1. Multimodal detection from routine hospital data

Single sources carry signal for both diseases. Chest-radiograph models detect malignant pulmonary nodules [6] and identify people at high risk of lung cancer [7], routine blood counts and clinical data flag lung cancer before diagnosis [8], and the ECG can identify left ventricular systolic dysfunction [9]. Multimodal models that fuse imaging with electronic health records (EHR) generally do better than single-source models [10,11]. On MIMIC-IV, the Holistic AI in Medicine (HAIM) framework fused tabular data, time series, notes and radiographs and used Shapley values to estimate each modality’s contribution [12]. Adding radiographs to clinical parameters improved the area under the receiver operating characteristic curve (AU-ROC) [13], and architectures for missing or inconsistent modalities have been proposed [14–16]. In the *Journal of Biomedical Informatics*, fusing structured data with notes improved prediction of acute kidney injury [17], and Integrated Gradients has been used to describe cross-modal contributions [18]. Yet the gains are not universal. In one fusion benchmark, fusion helped most where the evidence was spread across modalities and lost its advantage when inputs were missing [19], and elsewhere structured EHR alone matched multimodal inputs for several tasks [20]. The data that matter are therefore likely to differ by disease, and should be measured, not assumed.

### 2.2. Explanations, fairness and federated learning

Three further obstacles stand between such models and practice. The first is explanation. Saliency maps can look plausible while reflecting little of what the model uses, and post-hoc explanations often fail localisation and randomisation tests [21–23]; the randomisation test itself has known shortcomings [24], so no single test should decide validity. The second is fairness: chest-radiograph models can underperform in under-served groups [25], and methods to reduce demographic disparities in EHR models, including under class imbalance, have been proposed [26,27]. Performance must therefore be reported by sex, age, race and ethnicity, and socioeconomic position, as TRIPOD+AI requires [5]. Finally, hospitals rarely pool records, which makes federated learning attractive [28,29]. It has been benchmarked on healthcare datasets split naturally by centre [30] and evaluated across real data sources [31,32], and methods for missing modalities have begun to appear [33]. These questions have mostly been studied separately; few studies evaluate together, in one leakage-controlled framework, what each source contributes, whether explanations are valid for each source, how performance differs between subgroups, and what federated training costs relative to pooling.

## 3. Methods

### 3.1. Study design and data sources

This retrospective study develops and internally validates models that detect lung cancer and heart failure during a hospital stay from data recorded in the first 48 hours; Fig. 1 summarises the cohort, model, federated simulation and evaluation. The cohort definitions, models, hyper-parameters and primary hypothesis were fixed in a dated code release before data were accessed and were not tuned; analyses added later are listed in Supplementary Section S1.15, and full details are in Supplementary Methods S1, with the fixed settings in Supplementary Table S7 and the models compared in Supplementary Table S8. We used four linked PhysioNet [34] MIMIC databases from Beth Israel Deaconess Medical Center (BIDMC), Boston: MIMIC-IV version 3.1 [35,36], covering patients admitted through the emergency department (ED) or to intensive care between 2008 and 2022; MIMIC-CXR-JPG version 2.1.0 [37,38], chest radiographs from 2011–2016; MIMIC-CXR version 2.1.0 [39,40], the corresponding radiology reports; and MIMIC-IV-ECG version 1.0 [41], 10-second 12-lead ECGs from 2008–2019. The databases share patient identifiers and a common date shift, so records can be linked by patient and time.

**Figure 1:**
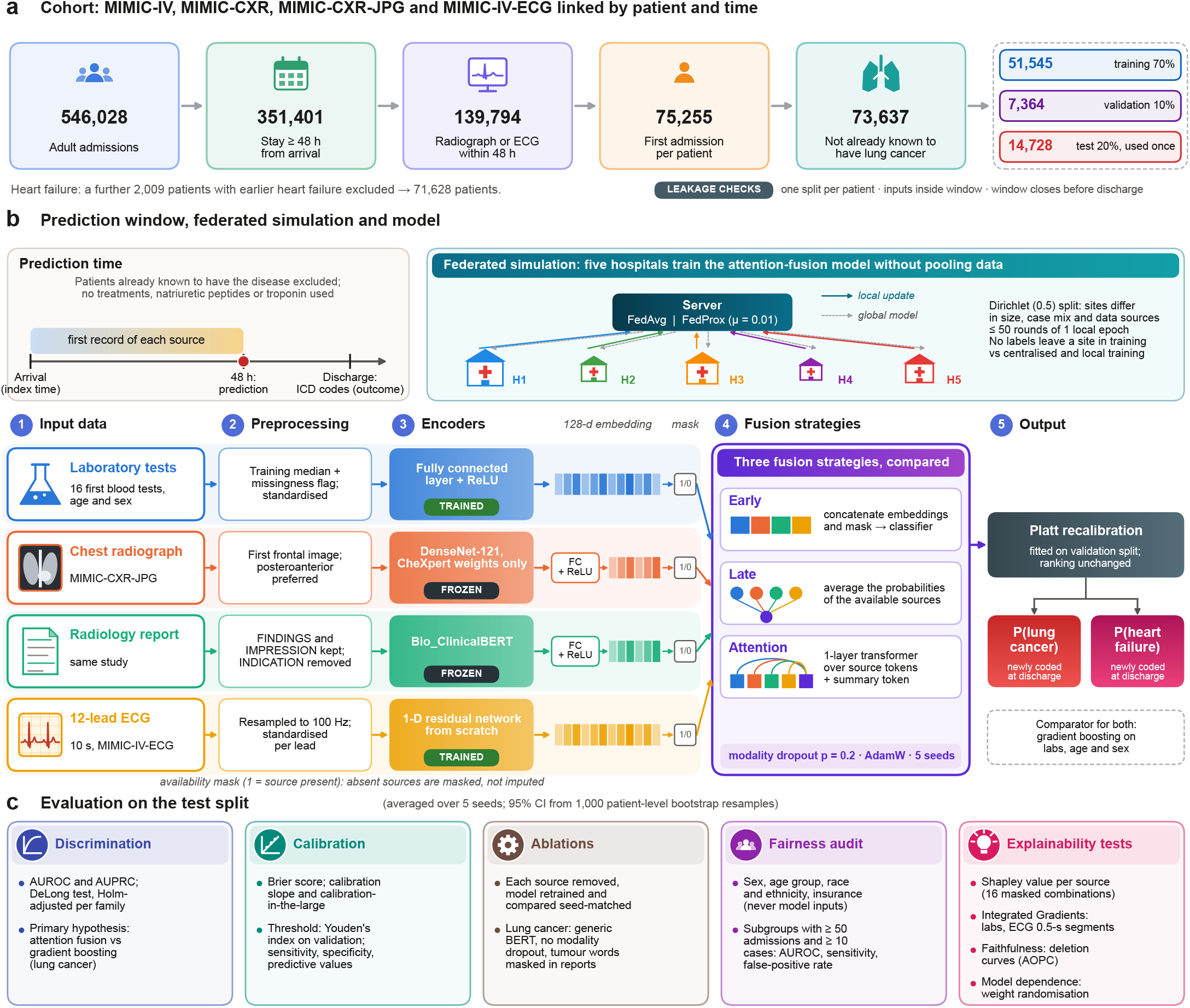
Study design and model architecture. (a) Cohort flow from MIMIC-IV admissions to the analysed cohort and its patient-level split. (b) Prediction window; federated simulation across five hospitals (FedAvg and FedProx); the four source encoders (frozen or trained), 128-dimensional embeddings and availability mask; and the three fusion strategies with Platt recalibration and the gradient-boosting comparator. (c) Evaluation on the test split.

### 3.2. Cohort and outcomes

The index time was hospital arrival: ED registration for patients who came through the ED, otherwise the admission time. Admissions were eligible if the patient was 18 years or older, the stay lasted at least 48 hours from the index time (so that a prediction at 48 hours precedes discharge), and at least one frontal chest radiograph or ECG was recorded within 48 hours of the index time. Only the first eligible admission of each patient was kept (Fig. 1a).

The primary outcome was lung cancer (International Classification of Diseases, Ninth and Tenth Revisions, Clinical Modification: ICD-9-CM 162.x; ICD-10-CM C33 or C34) and the secondary outcome heart failure (ICD-9-CM 428.x; ICD-10-CM I50.x), each in any position among the discharge diagnoses. To reduce the inclusion of disease already known, patients known to have lung cancer were excluded from the cohort: those with lung cancer coded in an earlier admission, or with a code for a personal history of lung cancer or for an encounter for radiotherapy, chemotherapy or immunotherapy in the index or an earlier admission. Patients with heart failure coded in an earlier admission were excluded from the heart-failure analyses. Outcomes were taken from discharge coding, so no outcome assessor interpreted the predictors.

### 3.3. Predictors

Every predictor had to be time-stamped within 48 hours of the index time; for each source the first record was used. No treatment given during the window was used as a predictor. **Laboratory tests:** age, sex and the first value of 16 routine blood tests (Supplementary Table S6), matched by patient and time so that ED samples were included. Natriuretic peptides and troponin were excluded because they are ordered when heart failure is already suspected. Missing values were replaced by the training median with a missingness indicator, and variables were standardised on the training split. **Chest radiograph:** the first frontal (posteroanterior preferred) image, encoded by a frozen DenseNet-121 [42] pretrained on CheXpert only [43,44], so that the encoder had never seen MIMIC-CXR. **Radiology report:** the FINDINGS and IMPRESSION sections of the same study, encoded by frozen Bio_ClinicalBERT [45]; the INDICATION section was removed because it records why the radiograph was ordered (for example, “? mass”). **ECG:** the first 12-lead ECG, resampled to 100 Hz and standardised per lead. Absent sources were not imputed; each model received an availability mask.

### 3.4. Model architecture

Each source was mapped to a 128-dimensional embedding: a single trained fully connected layer with ReLU activation for the EHR and for the frozen image and text embeddings, and a one-dimensional residual network [46] trained from scratch for the raw ECG (Fig. 1b). For each disease we compared single-source models; gradient boosting on the EHR features [47] as a strong conventional baseline; early fusion (concatenated embeddings and availability mask); late fusion (average of the single-source probabilities over available sources); and attention fusion, in which a one-layer transformer [48] attends over the available source embeddings and a learned summary token, with absent sources masked. Fusion models were trained with modality dropout (probability 0.2) [49]. Neural models were trained with AdamW [50], class-weighted binary crossentropy and early stopping on validation AUROC, each with five random seeds. Because class weighting inflates predicted risks, every model was recalibrated by Platt scaling [51], a logistic regression on the model’s logit fitted on the validation split separately for each seed; this changes calibration but not the ranking of patients, so AUROC is unaffected. Ablations removed each source in turn for both diseases; for lung cancer they also replaced Bio_ClinicalBERT with generic BERT [52] and removed modality dropout. All ablations were retrained from scratch, mostly with the first two of the five seeds because of the computing budget, and each was compared with the full model averaged over the same seeds.

#### Report masking check

Because a report may name a suspected tumour, the report model and attention fusion were retrained with 20 tumour-related word patterns (for example “mass”, “nodule”, “malignan*”, “carcinoma”; full list in Supplementary Methods S1) deleted from every report.

### 3.5. Data splitting, evaluation and statistical analysis

Patients were split into training (70%), validation (10%) and test (20%) sets stratified by lung cancer; the same split was used for heart failure. A script verified that no patient appears twice or in two splits, that every input lies inside the window and that the window closes before discharge. Preprocessing parameters were fitted on the training split; the validation split selected the epoch and the decision threshold (Youden’s index [53]); the test split was used for the final evaluation. A one-seed trial run that checked the pipeline end to end also scored the test split once; model settings were not changed afterwards (Supplementary Section S1.15).

The prediction of each model is the average over its five seeds. We report AUROC, area under the precision–recall curve (AUPRC), Brier score, calibration plots, calibration-in-the-large and calibration slope [54], and sensitivity, specificity and predictive values at the validation threshold, with 95% confidence intervals (CIs) from 1,000 patient-level bootstrap resamples, which reflect test-sample variability with the trained models held fixed; differences were computed from unrounded estimates. We also report sensitivity and positive predictive value at thresholds that flag fixed shares of validation patients, net benefit by decision-curve analysis [55], and performance by arrival route, radiograph availability and position of the outcome code (Supplementary Section S1.16). No a priori sample size calculation was performed because all eligible admissions were used; the number of events in each split and the precision of every estimate are reported [56]. AUROCs were compared with DeLong’s test [57,58]. The single pre-specified primary hypothesis was that attention fusion detects lung cancer better than gradient boosting on all test admissions, tested at a two-sided alpha of 0.05. All other tests, including all heart-failure tests, were grouped into families (model comparisons, ablations, the report masking check, federated comparisons and each fairness attribute, for each disease) and Holm-adjusted within each family [59]. Comparisons with a single-source model were made on the test admissions that had that source.

#### Fairness

Performance was evaluated by sex, age group (18–49, 50–64, 65–79 and 80 years or older), race and ethnicity (White, Black, Hispanic/Latino, Asian, other, unknown, as recorded in MIMIC-IV) and insurance as a socioeconomic proxy. Sex was taken as recorded in MIMIC-IV (female or male); gender identity is not recorded and was not studied. Race, ethnicity and insurance were never model inputs. For each subgroup with at least 50 test admissions and 10 cases we report AUROC, AUPRC, Brier score, calibration-in-the-large, and sensitivity and false-positive rate at the single validation threshold, so that differences correspond to unequal opportunity and unequal false alarms. Each subgroup was compared with the largest subgroup of the same attribute.

### 3.6. Federated learning simulation

Because MIMIC-IV comes from one hospital, we simulated five hospitals by spreading each combination of admission source and admission type over the sites with Dirichlet (0.5) proportions [60], repeating the draw until every site held at least 5% of the training data and 10 cases in its training and test parts. We compared centralised training on pooled data, local training at each site, federated averaging (FedAvg) [29] and FedProx [61], which adds a proximal term (*μ* = 0.01) that limits how far each site’s model drifts from the shared one, all using the attention-fusion model and the patient-level splits above, with up to 50 rounds of one local epoch and three seeds. The round kept was the one with the best size-weighted mean of per-site validation AUROCs, so no labels left a site during training; Platt recalibration of the federated models used the pooled validation split (Supplementary Section S1.15). The main summary is the share of the centralised gain recovered, *R* = (AUROC_federated_ −AUROC_local_)/(AUROC_centralised_ − AUROC_local_), with a bootstrap confidence interval. The simulation includes no differential privacy or secure aggregation [62], so no formal privacy guarantee is claimed.

### 3.7. Explainability evaluation

All explanations were computed for the trained attention-fusion model of each disease. On the whole test split, each admission was re-scored with all 16 combinations of its four sources masked, giving the AUROC lost when a source is removed and the Shapley value [63] of each source for AUROC and for each admission’s prediction; these were averaged over the five seeds. Within sources, Integrated Gradients [64] attributed the prediction to laboratory features (128 steps) and to 0.5-second segments of each ECG lead (32 steps), on 1,000 randomly drawn test admissions with an ECG and the first seed. Faithfulness was tested with deletion curves [65]: the top 1 to 10 laboratory features, or up to half of the ECG segments, were removed in order of attribution and compared with random removal (five random orders) by the area over the perturbation curve (AOPC). Model dependence was tested by cascading randomisation of the weights from the prediction head down to the input layers [21]; an explanation failed if the rank correlation of its absolute attributions with the original stayed at 0.5 or above after all layers were randomised. We also report the correlation of signed attributions. The deletion test reset features to the same baseline that Integrated Gradients used.

### 3.8. Ethics, data protection and code

The MIMIC databases are de-identified; their creation and sharing were approved by the institutional review boards of BIDMC and the Massachusetts Institute of Technology with a waiver of informed consent [35]. Every author who handled patient-level data held PhysioNet credentialed access and signed the data use agreements. Separate ethics approval was not required by the University of Dhaka for this secondary analysis of de-identified, publicly available data. All patient-level processing took place on the analysing author’s computer; only aggregate results left it, counts below 10 are suppressed in every reported table and figure, and no patient-level data were sent to any online service. There was no patient or public involvement. The analysis plan was fixed in a dated code release before data access; the code is described under Code availability.

## 4. Results

### 4.1. Study cohort

Of the 546,028 admissions in MIMIC-IV (all in adults), 351,401 lasted at least 48 hours from arrival and 139,794 of these had a chest radiograph or ECG within the first 48 hours. Keeping the first eligible admission of each patient left 75,255 admissions, and excluding patients already known to have lung cancer left 73,637 patients (Fig. 1a). They were split into 51,545 training, 7,364 validation and 14,728 test patients. Of the cohort, 76.9% arrived through the ED; the median age was 66 years (interquartile range (IQR) 53–78) and 49.0% were female.

Lung cancer was coded in 1,263 patients (1.72%): 884 in the training, 126 in the validation and 253 in the test split. For the heart-failure analyses, 2,009 patients with heart failure coded in an earlier admission were excluded, leaving 71,628 patients, of whom 15.6% had heart failure coded (7,781 of 50,165 training, 1,122 of 7,162 validation and 2,295 of 14,301 test patients). Outcome rates and characteristics were similar across the three splits (Table 1).

**Table 1:**
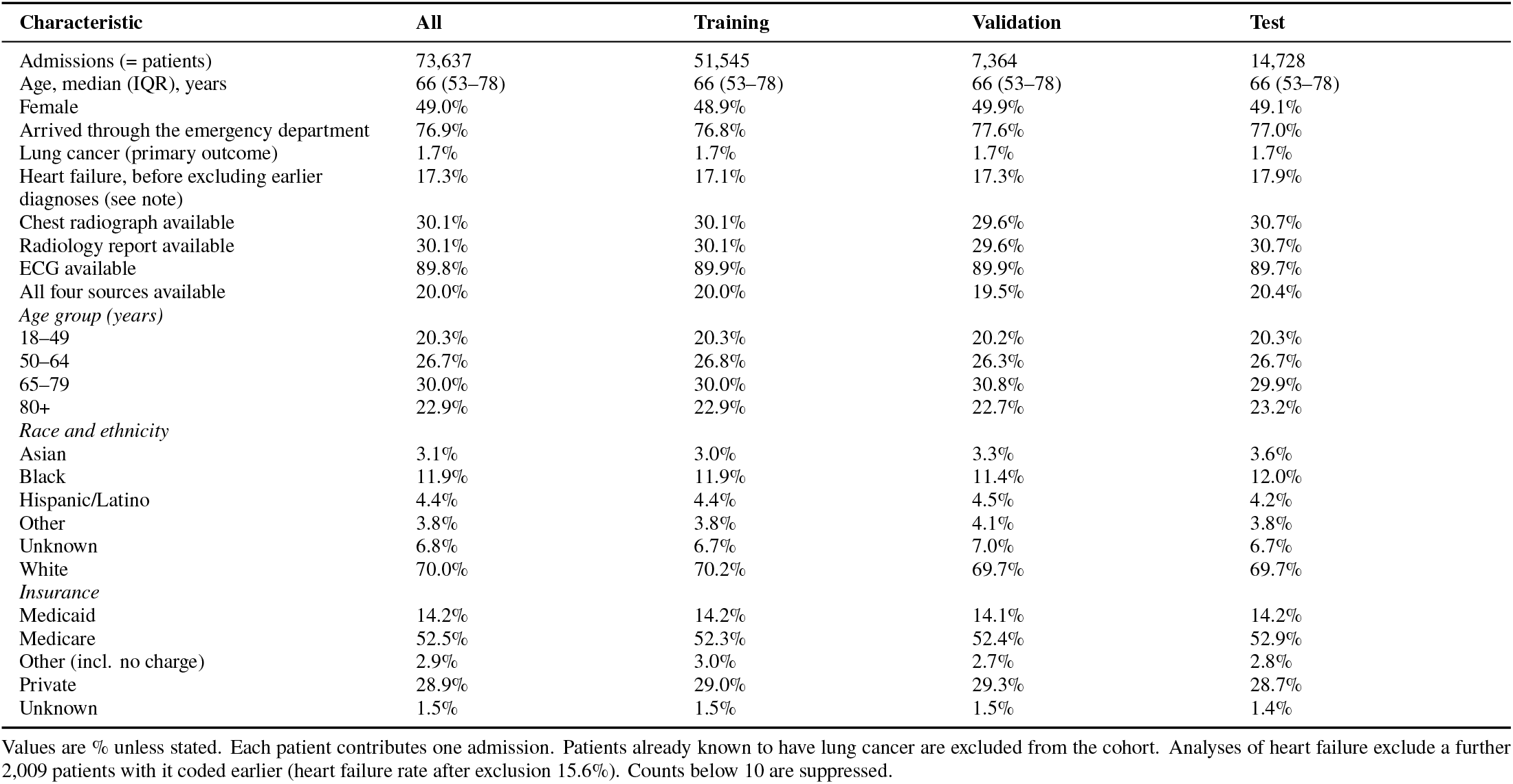
Characteristics of the study cohort by data split.

An ECG within 48 hours was available for 89.8% of patients, a chest radiograph with its report for 30.1%, and all four sources for 20.0%. Most routine laboratory tests were missing in fewer than 2% of patients; albumin was missing in 53.9% (Supplementary Table S6).

### 4.2. Detection of lung cancer

The primary hypothesis was supported. On all 14,728 test patients, attention fusion detected lung cancer with an AUROC of 0.833 (95% CI 0.809–0.859), against 0.753 (0.723–0.781) for gradient boosting on the laboratory tests, age and sex, a difference of 0.081 (DeLong p < 0.001; Table 2, Fig. 2a). The AUPRC, which must be read against a case rate of 1.7%, was 0.180 against 0.063. The gain did not depend on the fusion design. Early fusion reached 0.839 (0.815–0.864) and late fusion 0.829 (0.802–0.854), and attention fusion differed from neither (differences −0.006 and 0.005; Holm-adjusted p = 0.88 and p > 0.99; Supplementary Table S1).

**Table 2:** Detection of lung cancer and heart failure on the test set.

| Model | Test admissions (cases) | AUROC (95% CI) | AUPRC (95% CI) | Brier | Sensitivity | Specificity |
| --- | --- | --- | --- | --- | --- | --- |
| *Lung cancer* |  |  |  |  |  |  |
| Gradient boosting (labs, age, sex) | 14,728 (253) | 0.753<br>(0.723–0.781) | 0.063<br>(0.049–0.087) | 0.017 | 61.7% | 75.4% |
| Neural network (labs, age, sex) | 14,728 (253) | 0.754<br>(0.724–0.784) | 0.075<br>(0.055–0.106) | 0.017 | 70.0% | 68.0% |
| Chest radiograph | 4,516 (96) | 0.772<br>(0.723–0.818) | 0.098<br>(0.065–0.155) | 0.020 | 60.4% | 78.0% |
| Radiology report | 4,516 (96) | 0.890<br>(0.849–0.926) | 0.317<br>(0.224–0.426) | 0.017 | 78.1% | 85.2% |
| ECG | 13,212 (212) | 0.668<br>(0.631–0.703) | 0.037<br>(0.028–0.055) | 0.016 | 54.7% | 67.4% |
| Early fusion | 14,728 (253) | 0.839<br>(0.815–0.864) | 0.204<br>(0.159–0.261) | 0.015 | 70.4% | 80.0% |
| Late fusion | 14,728 (253) | 0.829<br>(0.802–0.854) | 0.179<br>(0.136–0.230) | 0.015 | 63.2% | 84.1% |
| Attention fusion | 14,728 (253) | 0.833<br>(0.809–0.859) | 0.180<br>(0.138–0.232) | 0.015 | 66.4% | 81.8% |
| *Heart failure* |  |  |  |  |  |  |
| Gradient boosting (labs, age, sex) | 14,301 (2,295) | 0.794<br>(0.785–0.804) | 0.432<br>(0.411–0.454) | 0.112 | 77.2% | 66.1% |
| Neural network (labs, age, sex) | 14,301 (2,295) | 0.801<br>(0.792–0.810) | 0.448<br>(0.428–0.470) | 0.111 | 81.7% | 62.5% |
| Chest radiograph | 4,376 (723) | 0.832<br>(0.816–0.846) | 0.519<br>(0.481–0.557) | 0.105 | 70.3% | 78.4% |
| Radiology report | 4,376 (723) | 0.811<br>(0.795–0.828) | 0.520<br>(0.480–0.560) | 0.107 | 67.5% | 79.7% |
| ECG | 12,849 (2,172) | 0.854<br>(0.846–0.862) | 0.568<br>(0.547–0.591) | 0.102 | 81.0% | 74.6% |
| Early fusion | 14,301 (2,295) | 0.886<br>(0.879–0.892) | 0.636<br>(0.615–0.656) | 0.088 | 82.8% | 77.6% |
| Late fusion | 14,301 (2,295) | 0.881<br>(0.875–0.888) | 0.621<br>(0.601–0.642) | 0.090 | 84.8% | 74.6% |
| Attention fusion | 14,301 (2,295) | 0.885<br>(0.878–0.892) | 0.637<br>(0.616–0.658) | 0.088 | 84.7% | 75.4% |

Seed-averaged predictions of 5 training runs; 95% CIs from 1,000 patient-level bootstrap resamples. Sensitivity and specificity at the Youden threshold chosen on the validation set. Single-source models are scored on test admissions that have that source; fusion and EHR models on all test admissions. Heart-failure analyses exclude patients with heart failure coded in an earlier admission.

**Figure 2:**
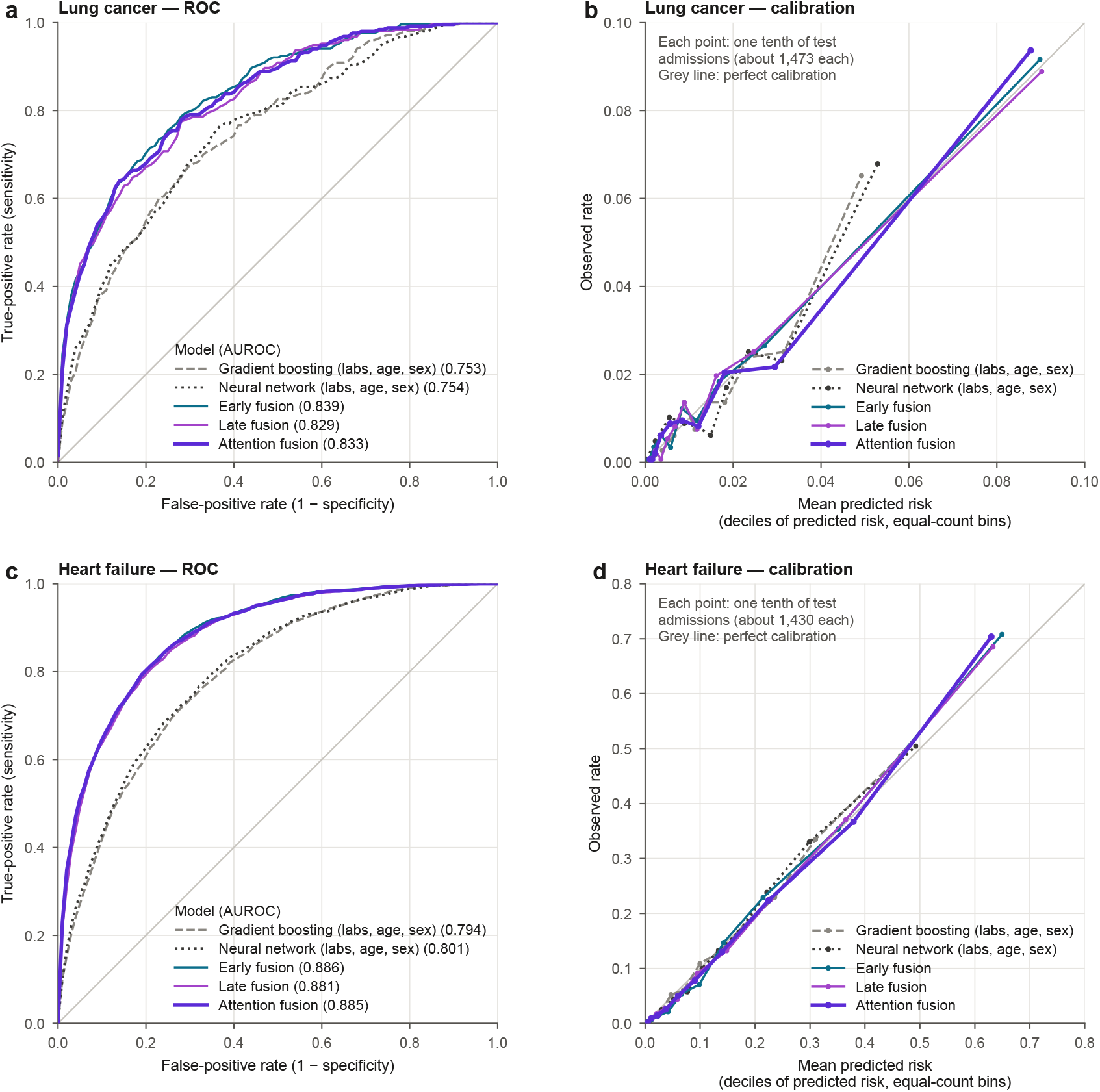
Test-set discrimination and calibration. (a, b) Lung cancer: receiver operating characteristic curves of gradient boosting, the neural EHR model and the three fusion models, and calibration by deciles of predicted risk. (c, d) The same for heart failure.

Among single sources, the radiology report was by far the strongest, with an AUROC of 0.890 (0.849–0.926) on the 4,516 test patients who had a report, against 0.772 for the radiograph image in the same patients, 0.754 for the laboratory network on all patients and 0.668 for the ECG on the 13,212 patients who had one. In these patients attention fusion reached 0.902, no different from the report alone (difference 0.012, adjusted p = 0.88) but well above gradient boosting (0.737); in the 10,212 patients without a radiograph its advantage over gradient boosting was 0.027 (unadjusted p = 0.087; Supplementary Table S11). In the 3,000 test patients with all four sources, the AUROC was 0.905 for attention fusion, 0.912 for early fusion, 0.902 for late fusion and 0.720 for gradient boosting.

Before recalibration, class weighting made the early- and attention-fusion models overestimate risk (calibration-in-the-large 0.29 and Brier score 0.142 for attention fusion). After Platt scaling, calibration-in-the-large was close to zero for every model, the calibration slope of attention fusion was 1.05 and its Brier score 0.0155 (Fig. 2b). At the threshold chosen on the validation split, attention fusion had a sensitivity of 66.4%, a specificity of 81.8%, a positive predictive value of 6.0% and a negative predictive value of 99.3%.

#### Report masking

Deleting the tumour-related words lowered the AUROC of the report model from 0.890 to 0.881 (difference 0.009, Holm-adjusted p = 0.036) and that of attention fusion from 0.833 to 0.818 (difference 0.015, Holm-adjusted p = 0.016). Most of the signal survived, but a report can describe a lesion in other terms, so the task is the detection of disease already visible in the first 48 hours, not the prediction of a future diagnosis.

#### Case mix and operating points

Of the 253 test cases, 45.1% had lung cancer coded as the principal diagnosis and 73.5% arrived through the ED. Attention fusion outperformed gradient boosting in ED arrivals (0.817 against 0.722) and when cases were restricted to principal (0.864 against 0.785) or secondary (0.808 against 0.726) coding (Supplementary Table S11). At a threshold that flagged 5% of validation patients, attention fusion detected 40.3% of test cases with a positive predictive value of 13.7%, against 21.3% and 7.8% for gradient boosting, and its net benefit exceeded that of gradient boosting and of treating everyone at threshold probabilities from 0.5% to 10% (Supplementary Tables S10 and S12).

### 4.3. Detection of heart failure

Heart failure was detected more accurately (Table 2, Fig. 2c–d). On all 14,301 test patients, attention fusion reached an AUROC of 0.885 (0.878–0.892) against 0.794 (0.785–0.804) for gradient boosting (difference 0.090, adjusted p < 0.001), with an AUPRC of 0.637 against 0.432. Early fusion reached 0.886 (0.879–0.892) and did not differ from attention fusion (adjusted p = 0.91); late fusion was slightly lower at 0.881 (difference 0.004, adjusted p = 0.009).

Here the ECG was the strongest single source, with an AU-ROC of 0.854 (0.846–0.862) on the 12,849 test patients who had one, followed by the radiograph image (0.832) and the report (0.811) on the 4,376 patients with a radiograph, and the laboratory network (0.801). In patients with an ECG, attention fusion improved on the ECG alone by 0.028 (adjusted p < 0.001). In the 2,924 test patients with all four sources, the AUROC was 0.894 for attention fusion, 0.895 for early fusion, 0.887 for late fusion and 0.781 for gradient boosting.

Recalibration again removed the overestimation caused by class weighting (calibration-in-the-large from 0.25 to close to zero; Brier score from 0.172 to 0.088). The calibration slope of attention fusion was 1.17, so its predictions were slightly less extreme than they should be (Fig. 2d). At the validation threshold, attention fusion had a sensitivity of 84.7%, a specificity of 75.4%, a positive predictive value of 39.7% and a negative predictive value of 96.3%. Heart failure was the principal diagnosis in 13.8% of test cases; attention fusion outperformed gradient boosting in ED arrivals (0.888 against 0.799) and in cases coded as principal (0.933 against 0.828) or secondary (0.877 against 0.789) diagnoses. When 10% of validation patients were flagged, 69.0% of flagged test patients had heart failure, against 47.9% with gradient boosting (Supplementary Tables S10–S12).

### 4.4. Which data matter for each disease

The two diseases relied on different sources (Table 3, Fig. 3a). Compared with the full model averaged over the same seeds, lung-cancer detection lost 0.080 AUROC without the laboratory data (adjusted p < 0.001) and 0.045 without the report (adjusted p = 0.002). Removing the ECG (0.011) or the radiograph image (0.002), replacing Bio_ClinicalBERT with generic BERT and training without modality dropout made no significant difference. For heart failure the ECG mattered most: removing it lost 0.058 AUROC and removing the laboratory data 0.020 (both adjusted p < 0.001); the radiograph image added 0.004 (adjusted p = 0.011) and the report nothing (adjusted p = 0.78).

**Table 3:**
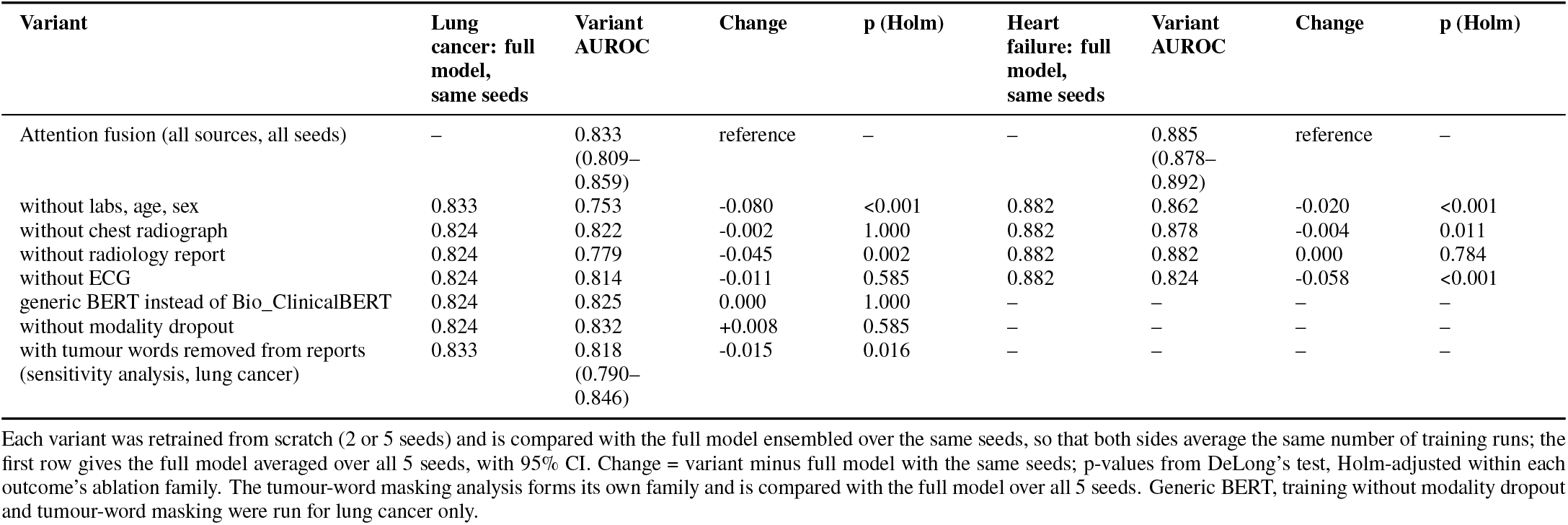
Ablation and sensitivity analyses of the attention-fusion model (test set).

**Figure 3:**
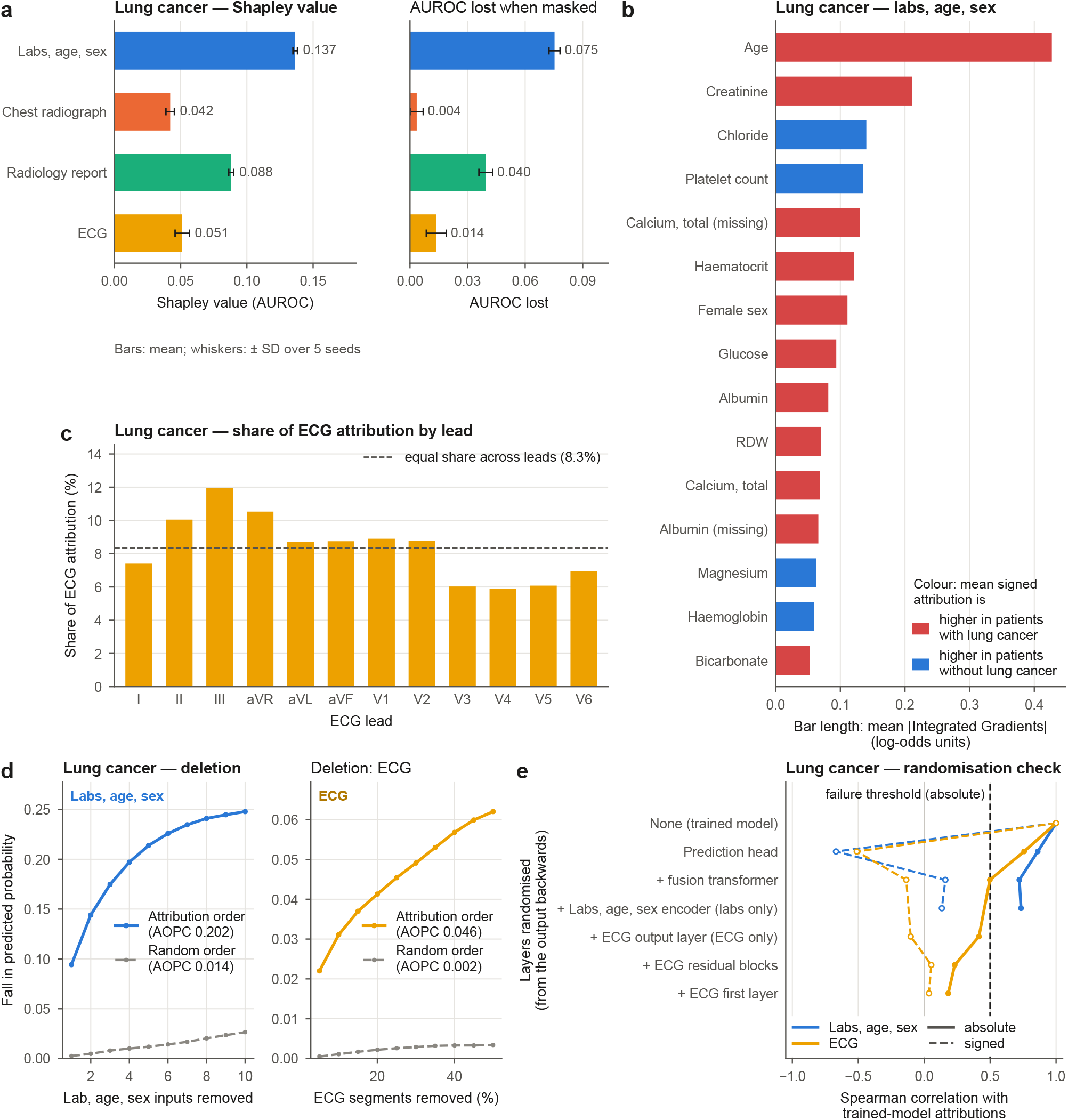
Explanations and their evaluation for lung cancer (attention-fusion model, test set). (a) Shapley value of each source for AUROC and AUROC lost when the source is masked (mean and SD over five seeds); computed per seed, the Shapley values sum to the mean single-seed AUROC minus 0.5, not to the AUROC of the five-seed ensemble. (b) Laboratory, age and sex attributions (Integrated Gradients). (c) Share of ECG attribution by lead. (d) Deletion curves: fall in predicted-class probability when features or ECG segments are removed in attribution order or at random. (e) Sanity check: rank correlation of absolute (solid) and signed (dashed) attributions with the original as layers are randomised from the output backwards; horizontal line, failure threshold. The same analyses for heart failure are in Supplementary Fig. S1.

Masking sources in the trained models without retraining gave a similar picture. For lung cancer, the Shapley values of the sources for AUROC were 0.14 for the laboratory data, 0.09 for the report, 0.05 for the ECG and 0.04 for the radiograph image; for heart failure they were 0.17 for the ECG, 0.13 for the laboratory data, 0.04 for the radiograph image and 0.03 for the report. The laboratory data made the largest contribution to the individual lung-cancer prediction in 52.0% of test patients, the ECG in 23.3%, the report in 21.7% and the radiograph image in 3.0%; for heart failure the ECG did so in 48.4% and the laboratory data in 35.8%. Where present, the report was the most influential source for lung cancer (mean absolute Shapley value 1.61 on the logit scale, against 1.01 for the laboratory data).

### 4.5. Are the explanations trustworthy?

Within the laboratory data, Integrated Gradients attributed most of the lung-cancer prediction to age (20% of the total attribution), creatinine (10%), chloride, platelet count and a missing calcium measurement; for heart failure, to age (15%), red-cell distribution width, urea nitrogen, creatinine and bicarbonate (Fig. 3b; Supplementary Fig. S1b). Attribution was spread across the ECG leads for lung cancer (5.9% to 11.9% per lead) and was highest in V1 (11.2%) for heart failure (Fig. 3c; Supplementary Fig. S1c).

The attributions passed the deletion test (Fig. 3d; Supplementary Fig. S1d). Removing laboratory features in order of attribution changed the predictions far more than removing them in random order: the AOPC difference was 0.188 (95% CI 0.180–0.196) for lung cancer and 0.134 (0.129–0.139) for heart failure, and the ordered removal did better than random removal in every one of the 1,000 explained patients for both diseases. For the ECG, the AOPC difference was 0.043 (0.041– 0.046) for lung cancer and 0.207 (0.201–0.212) for heart failure, with ordered removal better than random in 99.1% and 99.8% of patients.

On absolute attributions, the sanity checks separated the two sources (Fig. 3e; Supplementary Fig. S1e). After all layers were randomised, the rank correlation with the original fell to 0.18 for lung cancer and 0.16 for heart failure for the ECG, which passed, but stayed at 0.73 for the laboratory data, above the pre-specified limit of −0.5. On signed attributions the laboratory correlations fell to 0.13 and 0.05. The absolute ranking of laboratory features therefore reflects how far each input lies from the baseline at least as much as what the model learned, even though the deletion test shows that the highly ranked features do move the trained model’s predictions. Only 21 of the 1,000 patients explained for lung cancer had the disease, so the feature-level explanations for lung cancer describe mainly patients without it.

### 4.6. Fairness

For lung cancer (Fig. 4; Supplementary Tables S2–S3), attention fusion discriminated equally well in women and men (AUROC 0.833 and 0.834; adjusted p = 0.96), although at the shared threshold its sensitivity was higher in women (71.1% against 61.0%). The largest difference was in the youngest group. The AUROC was 0.950 in patients aged 18–49, who had only 11 cases, against 0.787 in those aged 65–79, the largest group (difference 0.163, 95% CI 0.106–0.219, adjusted p < 0.001); patients aged 50–64 and 80 or older did not differ significantly from those aged 65–79. No racial or ethnic group scored significantly lower than White patients (0.816): the AU-ROC was 0.892 for Black patients and 0.896 for patients of unknown race (adjusted p = 0.12 for both), while Asian and Hispanic/Latino patients had fewer than 10 cases and could not be scored. By insurance, patients with Medicaid (0.877) or private insurance (0.871) scored higher than those with Medicare (0.789; adjusted p = 0.012 for both).

**Figure 4:**
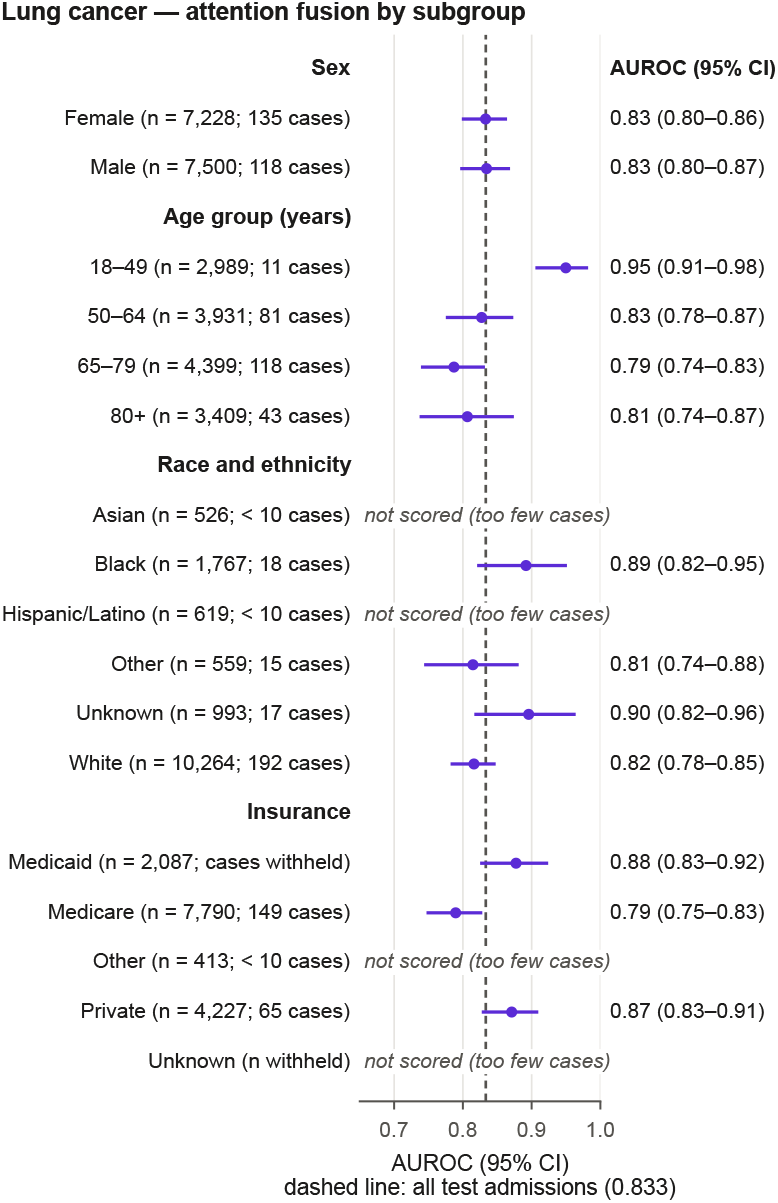
AUROC (95% CI) of the attention-fusion model for lung cancer by sex, age group, race and ethnicity, and insurance. Dashed line, all test admissions. Subgroups with fewer than 50 test admissions or 10 cases are not scored; categories with fewer than 10 test admissions are not shown, and the size or case count of the smallest shown categories of that attribute is withheld so that suppressed counts cannot be derived.

For heart failure (Supplementary Table S5), women and men again did not differ (0.884 and 0.886; adjusted p = 0.84). Discrimination fell with age, from 0.911 in patients aged 18–49 to 0.812 in those aged 80 or older (80 or older against 65– 79: difference −0.046, 95% CI −0.068 to −0.025, adjusted p < 0.001). At the shared threshold this produced very different trade-offs: in patients aged 80 or older the sensitivity was 91.5% and the false-positive rate 55.2%, against 60.4% and 4.7% in those aged 18–49, whose case rates were 29.2% and 3.6%. Hispanic/Latino patients scored higher than White patients (0.929 against 0.878, adjusted p = 0.005), and no group scored significantly lower. Patients with Medicare scored lower (0.850) than those with Medicaid (0.904) or private insurance (0.905; adjusted p < 0.001 for both). In every scored subgroup of both diseases, calibration-in-the-large stayed close to zero in absolute terms, although with lung-cancer case rates of a few per cent small absolute errors can mean large relative ones.

### 4.7. Federated training

The five simulated sites differed in size (4,464 to 24,242 training patients for lung cancer) and case rate (1.5% to 2.2%; Supplementary Table S4). For lung cancer (Table 4), centralised training on the pooled data reached an AUROC of 0.830 (0.805– 0.858) with the three seeds used in this simulation, and local training at each site reached 0.784 (0.754–0.814), a gap of 0.046 (adjusted p < 0.001). FedAvg reached 0.807 (0.778–0.837), recovering 51% of that gap (95% CI 6% to 83%), and remained below centralised training (difference 0.023, adjusted p = 0.014). FedProx reached 0.822 (0.793–0.849), recovering 82% (51% to 112%); it was better than FedAvg (difference 0.014, adjusted p = 0.011) and did not differ significantly from centralised training (adjusted p = 0.20). FedAvg did not differ significantly from local training (adjusted p = 0.078), whereas FedProx did (adjusted p = 0.003). At the smallest site, with 17 test cases, the AUROC was 0.651 with local training, 0.740 with FedAvg and 0.760 with FedProx, with wide intervals.

**Table 4:** Federated training across simulated hospitals: lung cancer (attention-fusion model, test set).

| Training | AUROC (95% CI) | AUPRC (95% CI) | Brier | AUROC range across sites | Round kept (per seed) |
| --- | --- | --- | --- | --- | --- |
| Centralised (pooled data) | 0.830 (0.805–0.858) | 0.175 (0.133–0.229) | 0.016 | 0.743–0.902 | – |
| Local only | 0.784 (0.754–0.814) | 0.134 (0.100–0.180) | 0.016 | 0.651–0.843 | – |
| FedAvg | 0.807 (0.778–0.837) | 0.150 (0.113–0.198) | 0.016 | 0.740–0.861 | 27, 18, 41 |
| FedProx | 0.822 (0.793–0.849) | 0.163 (0.123–0.213) | 0.016 | 0.760–0.894 | 19, 29, 42 |

5 simulated hospitals (Dirichlet alpha 0.5); same patient-level test set for every method. Data exchanged per round: 28.0 MB. FedAvg recovered 50.8% of the centralised gain over local training (seed-averaged predictions; 95% CI 6.4% to 83.5%), and 74.2% when computed on the mean AUROC of the individual seeds. FedProx recovered 81.8% of the centralised gain over local training (seed-averaged predictions; 95% CI 51.2% to 112.5%), and 97.8% when computed on the mean AUROC of the individual seeds. FedAvg vs local only: AUROC difference +0.024, $p = 0.078$ (DeLong, Holm-adjusted within the federated family). FedProx vs local only: AUROC difference +0.038, $p = 0.003$ (DeLong, Holm-adjusted within the federated family).

For heart failure (Supplementary Table S9), the gap between centralised training (0.883) and local training (0.862) was smaller, 0.022. FedAvg and FedProx both reached 0.882, recovering 95% of the gap (95% CI 86% to 105% for FedAvg), and neither differed from centralised training (adjusted p = 0.72) or from the other. Federated training reached 95% of its best validation gain within 18 rounds and exchanged 28.0 MB of model weights per round (Supplementary Fig. S2).

## 5. Discussion

### 5.1. Principal findings

Across 73,637 patients, fusing laboratory tests, the ECG, the chest radiograph and its report from the first 48 hours detected lung cancer coded at discharge with an AUROC of 0.833 and heart failure with 0.885, gains of 0.081 and 0.090 over gradient boosting on laboratory data. The fusion design mattered little; the sources mattered a great deal. Lung cancer depended on the laboratory data and, where present, the report, and heart failure on the ECG and the laboratory data. Explanations passed a deletion test for both sources, but their verdict in the randomisation check depended on the source and the similarity measure. Discrimination was equal by sex and, for heart failure, fell with age. Simulated federated training kept most of the pooled benefit for heart failure, and for lung cancer more with FedProx than with FedAvg.

### 5.2. Comparison with previous work

Unlike curated imaging studies [6,7], we studied unselected admissions, so accuracies are not directly comparable. The disease-specific reliance on sources agrees with single-source work: the ECG, the main heart-failure source here, identifies left ventricular dysfunction on its own [9], and routine laboratory data carry early signal of lung cancer [8]. HAIM also used Shapley values for modality contributions [12]; we add per-source tests of the explanations, a fairness audit and a federated comparison. That the choice of sources mattered more than the fusion design agrees with benchmarks of EHR and radiograph fusion on MIMIC [19,20], and, as in earlier work [13], adding sources improved discrimination over the EHR alone. Simple early fusion with an availability mask did as well as attention fusion, even though only 20.0% of patients had all four sources. We did not re-implement architectures built for missing modalities [14–16], so our results speak to which data matter rather than to state-of-the-art accuracy.

### 5.3. Explanations

The deletion tests showed that the highest-ranked features moved the predictions for both diseases and both sources. The laboratory attributions, however, kept their absolute ranking when all the weights were randomised, the failure described by Adebayo et al. [21], although their signed ranking changed. Integrated Gradients multiplies the gradient by the difference between input and baseline, so for the single layer that encodes the laboratory data the absolute ranking is dominated by how far each value lies from the baseline; randomisation checks are known to be weak for shallow encoders and to depend on the similarity measure [24]. Our deletion test also used the Integrated Gradients baseline, which favours that method. The ECG attributions, computed through a deep convolutional encoder, passed both tests. Saliency methods often look plausible while failing such tests [22,23]; our results add that, within one model, the verdict can differ between sources and measures, so explanations should be tested per source and with more than one measure before they are displayed. At the level of sources, retrained ablations and masking of the trained model broadly agreed.

### 5.4. Fairness

Discrimination did not differ by sex, and no racial or ethnic group had significantly lower discrimination than White patients, but Asian and Hispanic/Latino patients had too few lung-cancer cases to be assessed and several intervals were wide. We therefore did not see the underdiagnosis reported for chest-radiograph models [25], although small groups limit this conclusion. The clearest gradient was age in heart failure, where coexisting disease in older patients makes the condition harder to separate from other causes of abnormal tests; the insurance differences may largely mirror age. A shared threshold behaved very differently across ages: for heart failure it flagged most older patients, many falsely, and missed many cases in younger patients. A hospital deploying such a model would need to monitor alert rates, sensitivity and calibration by age group, and could consider age-specific thresholds.

### 5.5. Federated learning

With data from one centre, federated training cannot be expected to beat centralised training; the question is how much of the benefit of pooling it keeps. For heart failure, a common outcome, both algorithms kept almost all of it. For lung cancer, with as few as 68 training cases at the smallest site, FedAvg kept about half and did not differ significantly from local training, whereas FedProx, which limits how far each site’s model drifts from the shared one, kept most of it. Both shares were higher for single-seed models and come from one simulated partition and one value of *μ*, so they suggest rather than show that drift control matters for rare outcomes; benchmarks with natural multi-site splits [30] are needed. Real differences between hospitals in coding, equipment and populations are likely to be larger.

### 5.6. Strengths and limitations

The main strengths are the patient-level, leakage-checked design with every input time-stamped before the prediction time; the exclusion of patients already known to have the disease; an image encoder that never saw MIMIC-CXR; a pre-specified primary hypothesis with multiplicity control; explanations that were tested for faithfulness and model dependence rather than only shown; a fairness audit; and complete code with automated tests, to be released on publication.

The study has limitations. First, all data come from one academic centre, and the federated sites were simulated from that centre’s data; external and temporal validation are needed before clinical use. Second, outcomes came from discharge ICD codes, which miss some cases and include some not clinically confirmed, and the task is detection before discharge of disease coded at discharge, not prediction of a future diagnosis. Disease known from outpatient care or other hospitals cannot be seen in MIMIC-IV, and heart failure was the principal diagnosis in 13.8% of cases. A suspicious radiograph or report can prompt the work-up that leads to the code, which may inflate discrimination, particularly for the report, although results in ED arrivals and by diagnosis position were similar; 34 of the 253 lung-cancer test cases were same-day surgical admissions, in which the disease was probably known. Third, lung cancer is uncommon, which limits the precision of subgroup and per-site estimates. Fourth, the cohort includes only patients admitted through the ED or to intensive care who stayed at least 48 hours, and radiographs exist only for 2011–2016, so source availability is partly confounded with calendar period and the 2015 change from ICD-9 to ICD-10. Fifth, report sign-off times are not released; we assumed that each report reflects its radiograph as read soon after acquisition. Sixth, the image and text encoders were frozen and generic, so the small contribution of the radiograph applies to a frozen CheXpert encoder, and Bio_ClinicalBERT was pretrained on earlier notes from the same hospital. Seventh, missingness indicators can encode clinicians’ test-ordering behaviour (a missing calcium measurement ranked among the lung-cancer attributions). Eighth, the comparator used laboratory data only, and some analyses were added after the protocol release (Supplementary Section S1.15). Finally, race and ethnicity are recorded administratively, and the explanations were not rated by clinicians.

### 5.7. Implications and future work

For lung cancer the most useful inputs were routine laboratory tests and the radiology report, both already in the record; for heart failure, the ECG. Most of the discrimination therefore came from data already stored, and this frozen image encoder added little. Because lung cancer is uncommon among admissions, even a good model flags many patients without it: at the validation threshold only 6.0% of flagged patients had lung cancer, whereas flagging 5% of patients found 40.3% of cases with a positive predictive value of 13.7%. Such output would suit a prompt to review existing imaging and arrange follow-up, not a diagnosis, and because the model largely detects disease already visible in the first 48 hours, its value would lie in ensuring such findings are acted on before discharge. Future work should validate the models externally and prospectively, measure the effect on time to diagnosis, and test federated training across real hospitals with secure aggregation.

## 6. Conclusion

Routinely collected laboratory tests, ECGs, chest radiographs and radiology reports from the first 48 hours of a hospital stay, used together, detected lung cancer and heart failure coded at discharge better than laboratory data alone, and the two diseases relied on different sources. Simple fusion was as good as attention-based fusion. Explanation checks gave different verdicts by source and similarity measure, and for heart failure discrimination fell with age. Simulated federated training kept most of the benefit of pooling for heart failure; for lung cancer, FedProx kept more than FedAvg. After external and prospective validation, such a model could run in the background to prompt review of patients at risk, and the evaluation framework, which tests explanations per source, audits fairness and compares federated with pooled training, can be reused for other diseases.

## Supporting information

TRIPOD+AI Checklist

## Data Availability

The data are available from PhysioNet to credentialed users who sign the respective data use agreements (MIMIC-IV, MIMIC-CXR, MIMIC-CXR-JPG and MIMIC-IV-ECG). Under these agreements the authors cannot share patient-level data. Aggregate results are included in the article and its Supplementary Material. The pre-specified analysis plan and code release v1.0-protocol are archived at https://doi.org/10.5281/zenodo.23506613.

https://doi.org/10.5281/zenodo.23506613

https://physionet.org/content/mimiciv/3.1/

https://physionet.org/content/mimic-cxr/2.1.0/

https://physionet.org/content/mimic-cxr-jpg/2.1.0/

https://physionet.org/content/mimic-iv-ecg/1.0/

## Declarations

### CRediT authorship contribution statement

Mostak Ahmed: Conceptualization, Methodology, Software, Formal analysis, Data curation, Visualization, Writing – original draft, Project administration. Md Mostafizur Rahman: Writing – review & editing. Sabera Tabassum Wahid: Writing – review & editing. Golam Rabbi: Writing – review & editing. Al Rafy: Writing – review & editing. Golam Sarwar Khondoker: Writing – review & editing. Shovit Dutta: Writing – review & editing. Mohammad Amir Saadat: Writing – review & editing. Md. Masuqul Haque: Supervision, Writing – review & editing.

### Data availability

The data are available from PhysioNet to credentialed users who sign the respective data use agreements: MIMIC-IV [36], MIMIC-CXR [40], MIMIC-CXR-JPG [38] and MIMIC-IV-ECG [41]. Under these agreements the authors cannot share patient-level data. Aggregate results are included in the article and its Supplementary Material.

### Code availability

The analysis plan, cohort definitions, model settings and automated tests were fixed in release v1.0-protocol, dated 5 October 2026, before the credentialed data were downloaded; this release is archived at Zenodo (https://doi.org/10.5281/zenodo.23506613). The complete pipeline, with automated tests on synthetic data, will be released publicly at https://github.com/mostakphoenixsoftbd/multimodal-lungcancer-hf when the article is published; until then it is available to editors and reviewers on request from the corresponding author.

### Ethics statement

See Section 3.8.

### Declaration of generative AI and AI-assisted technologies in the manuscript preparation process

During the preparation of this work the authors used generative AI tools (large language model-based assistants) in order to assist with programming the analysis code and with editing the language of the manuscript. The study design, data analysis and interpretation are the authors’ own; the tools received no patient-level data, and every number in the manuscript was produced by the authors’ scripts and checked automatically against the result files. No generative AI tool was used to create the figures. After using these tools, the authors reviewed and edited the content as needed and take full responsibility for the content of the published article.

## Acknowledgements

We thank the MIT Laboratory for Computational Physiology and BIDMC for making the MIMIC databases available.

