## Supplementary Material for "Explainable Multimodal AI for In-Hospital Detection of Lung Cancer and Heart Failure Using Routine Data: Development, Internal Validation, and Simulated Federated Evaluation"

#### Supplementary Methods S1. Full description of the methods

*This section gives the complete specification summarised in Section 3 of the main text. Every setting is the one in the released code (`config.py`).*

##### S1.1 Study design and reporting

This is a retrospective diagnostic (detection) modelling study that develops and internally validates models to detect two chronic diseases, lung cancer (primary outcome) and heart failure (secondary outcome), during a hospital admission from data recorded in the first 48 hours. It is reported according to the TRIPOD+AI statement [1]. The study definition, model list, hyper-parameters and analysis plan were fixed in code before any data were accessed, and none of them was tuned on the data; analyses added after the protocol release are listed in Section S1.15. All analyses were run on the credentialed author's own computer; only aggregate results left it (Section S1.13).

The study asks three questions: (i) whether combining routinely collected admission data from four sources detects each disease better than any single source; (ii) which sources and which features within them drive the predictions, and whether the explanations reflect what the model actually uses; and (iii) how much of the benefit of pooling data is kept when the model is trained by federated learning across hospitals that cannot share records.

##### S1.2 Data sources

Four linked PhysioNet databases [2] from the Beth Israel Deaconess Medical Center (BIDMC), Boston, were used:

- **MIMIC-IV** version 3.1 [3], hospital module (`hosp`): admissions, patients, discharge diagnoses (ICD-9-CM and ICD-10-CM) and laboratory results.
- **MIMIC-CXR-JPG** version 2.1.0 [4]: chest radiographs in JPEG format with study metadata (view position, study date and time).
- **MIMIC-CXR** version 2.1.0 [5]: the free-text radiology reports of the same studies.

- **MIMIC-IV-ECG** version 1.0 [6]: 10-second, 12-lead diagnostic electrocardiograms (ECGs) sampled at 500 Hz.

MIMIC-IV covers patients admitted through the emergency department (ED) or to intensive care at BIDMC between 2008 and 2022; MIMIC-CXR contains chest radiographs from 2011–2016 of patients seen in the ED; MIMIC-IV-ECG contains diagnostic ECGs from 2008–2019. The four databases share patient identifiers and a common de-identification date shift, so records can be linked by patient and time. Only the image and ECG files of the final cohort were downloaded.

##### S1.3 Cohort selection

The unit of analysis was the hospital admission. The index time, which starts the observation window, was hospital arrival: the ED registration time for patients who came through the ED (registration before the admission time), and otherwise the admission time. Many chest radiographs, ECGs and first blood tests are taken in the ED before formal admission, and MIMIC-CXR was assembled from ED radiographs, so starting the window at admission would discard them. Admissions were included if the patient was aged 18 years or older at admission (age = anchor age + admission year − anchor year; MIMIC-IV top-codes ages above 89), the stay lasted at least 48 hours from the index time, and at least one frontal chest radiograph (posteroanterior or anteroposterior) or one 12-lead ECG was recorded within 48 hours of the index time. The minimum length of stay guarantees that the window closes before discharge, so a prediction made at 48 hours is always made while the patient is still in hospital.

To reduce the inclusion of disease already known, patients known to have lung cancer were excluded from the cohort (Section S1.4). To keep observations independent, only the first eligible admission of each patient was kept. The cohort flow, with the number of admissions remaining after each step, is shown in main text Fig. 1a.

##### S1.4 Outcomes

The primary outcome was lung cancer, defined as any ICD-9-CM 162.x (malignant neoplasm of trachea, bronchus and lung) or ICD-10-CM C33 or C34 code, in any diagnostic position, among the discharge diagnoses of the index admission; secondary malignancy of the lung (metastases) was not counted. The second outcome was heart failure (ICD-9-CM 428.x, ICD-10-CM I50.x), defined in the same way. Patients already known to have lung cancer were excluded from the cohort: those with lung cancer coded in any earlier MIMIC-IV admission, or with a code in the index or any earlier admission for a personal history of lung cancer (ICD-9-CM V10.11; ICD-10-CM Z85.11) or for an encounter for radiotherapy, chemotherapy or immunotherapy (V58.0, V58.11, V58.12; Z51.0, Z51.11, Z51.12). Such codes show that the disease was diagnosed before the admission, possibly at another institution. Patients with heart failure coded in any earlier admission were excluded from the heart-failure analyses only; the splits, inputs and models were otherwise the same for both outcomes. Outcome codes are assigned at discharge by hospital coders and were never used as model inputs; no outcome assessor interpreted the predictors. "Not previously diagnosed" can only be judged within MIMIC-IV: a diagnosis recorded only at another institution and not reflected in these codes cannot be seen.

##### S1.5 Predictors

Every predictor had to be time-stamped within the window [index time, index time + 48 h). For each source, the first record in the window was used, including investigations done in the ED before admission. Treatments given during the window (medications, procedures) were not used as predictors.

**Structured EHR (labs, age, sex).** Age, sex and the first value of 16 routine blood tests: creatinine, urea nitrogen, sodium, potassium, chloride, bicarbonate, glucose, haemoglobin, haematocrit, white blood cells, platelet count, albumin, magnesium, total calcium, anion gap and red cell distribution width. Tests were matched by name in the MIMIC-IV laboratory dictionary and restricted to blood specimens. Results were matched to the admission by patient and time rather than by admission identifier, because samples taken in the ED before admission usually carry no admission identifier; since each patient contributes one admission, this assignment is unique. Natriuretic peptides and troponin were extracted but excluded from all models: they are usually ordered when a clinician already suspects heart failure, so they measure suspicion rather than early evidence. They were not used in any model. Missing laboratory values were replaced by the training-set median, a binary missingness indicator was added for each variable with any missing value, and all continuous variables were standardised with the training-set mean and standard deviation.

**Chest radiograph.** The first frontal radiograph in the window was used, with a posteroanterior view preferred when both views were taken at the same time. Images were reduced to 512 pixels on the shorter side when downloaded. For modelling, each image was converted to greyscale, centre-cropped to a square, resized to 224 × 224 pixels and scaled to the input range of TorchXRayVision [7]. A DenseNet-121 [8] pretrained by TorchXRayVision on CheXpert only [9] was used as a frozen encoder, giving a 1,024-dimensional embedding per image. Weights trained on MIMIC-CXR were deliberately avoided, because they could have seen this study's test images.

**Radiology report.** The report of the same radiograph study was used. Only the FINDINGS and IMPRESSION sections were kept; the INDICATION and HISTORY sections were removed because they record why the film was ordered (for example, "? CHF") and so measure clinician suspicion. De-identification placeholders were removed. Each report was encoded by Bio_ClinicalBERT [10] as a frozen encoder (maximum 512 tokens), and the last hidden layer was averaged over tokens to give a 768-dimensional embedding. Reports without either section (727 reports) were encoded as a fixed placeholder string. MIMIC-CXR does not release report sign-off times; the report is assumed to describe the radiograph as read at the time of the study.

**Electrocardiogram.** The first 12-lead ECG in the window was read with the WFDB software [11], arranged by lead name in a fixed order (I, II, III, aVR, aVF, aVL, V1–V6; figures show the leads in the conventional order), resampled from 500 Hz to 100 Hz (12 leads × 1,000 samples), standardised per lead and clipped at ±10 standard deviations. Unreadable records would have been treated as missing; none occurred. Unlike the image and text encoders, the ECG encoder was trained (Section S1.6).

**Missing sources.** The structured EHR input was available for every admission; the radiograph, report and ECG were each present or absent. Absent sources were not imputed. Each model received a binary availability mask and ignored absent sources (Section S1.6). In the test split, 30.7% of admissions had a radiograph and its report and 89.7% had an ECG (main text Table 1).

##### S1.6 Models

All neural models share one design (main text, Fig. 1b). Each source is first mapped to a 128-dimensional embedding by its own encoder:

- **EHR:** a linear layer with ReLU and dropout (0.2).

- **Radiograph and report:** layer normalisation of the frozen embedding, followed by a linear layer with ReLU and dropout (0.2).
- **ECG:** a one-dimensional residual network [12] trained from scratch: a convolutional stem (kernel 15, stride 2, 32 channels), four residual blocks (kernel 7, stride 2; 32, 64, 64 and 128 channels) with batch normalisation, global average pooling and a linear layer to 128 dimensions.

The embeddings are then combined by one of the following strategies (Supplementary Table S8):

- **Single-source models** use one encoder followed by a two-layer prediction head. A gradient-boosting classifier (scikit-learn `HistGradientBoostingClassifier` [13]; up to 500 iterations, balanced class weights, early stopping on 10% of the training split) on the same EHR features served as a strong conventional baseline.
- **Early fusion** concatenates the four embeddings (absent sources set to zero) with the availability mask and passes them to a two-layer prediction head.
- **Late fusion** averages the predicted probabilities of the four single-source models over the sources available for each admission.
- **Attention fusion** treats each available embedding as a token, adds a learned source-type embedding and prepends a learned summary token. One transformer encoder layer [14] (four attention heads, feed-forward width 256, dropout 0.2) mixes the tokens; absent sources are excluded through the attention padding mask. The prediction is read from the summary token through layer normalisation and a linear layer, so an admission always receives a prediction whatever sources it has.

Fusion models were trained with modality dropout [15]: during training, each available source was hidden with probability 0.2, never hiding all sources at once, so that the models learn to work with incomplete inputs.

**Supplementary Table S8. Models compared.**

| Group | Model | Inputs |
|---|---|---|
| Single source | Gradient boosting | Labs, age, sex |
| | Neural EHR | Labs, age, sex |
| | Radiograph | Frozen DenseNet-121 embedding |
| | Report | Frozen Bio_ClinicalBERT |

| Group | Model | Inputs |
|---|---|---|
| | | embedding |
| | ECG | Raw 12-lead signal, 1-D ResNet |
| Fusion | Early fusion | All four, concatenated |
| | Late fusion | Average of the four single-source probabilities |
| | Attention fusion (primary model) | All four, transformer over source tokens |
| Ablation of attention fusion | Without EHR / radiograph / report / ECG | Three of the four sources |
| | Generic text encoder | BERT-base [16] instead of Bio_ClinicalBERT |
| | No modality dropout | All four, dropout probability 0 |
| Report masking check | Report model and attention fusion | Reports with 20 tumour-related word patterns deleted |

**Training.** Neural models were trained with AdamW [17] (learning rate $10^{-3}$, weight decay $10^{-4}$, batch size 128) for up to 30 epochs on the training split, minimising binary cross-entropy with the positive class weighted by the ratio of negatives to positives in the training data. After every epoch the validation AUROC was computed; training stopped when it had not improved for five epochs, and the weights of the best epoch were kept. Single-source models were trained and evaluated only on admissions that had that source; fusion models used all admissions. Every primary model was trained with five random seeds (0–4); ablation and federated runs used fewer (Supplementary Table S7). The pretrained encoders were never fine-tuned, so the whole study runs on a laptop CPU without a graphics processor.

##### S1.7 Data splitting and leakage controls

Patients were randomly split into training (70%), validation (10%) and test (20%) sets, stratified by the primary outcome (random seed 42). Because each patient contributes one admission, the split is patient-level. The cohort script stops with an error if any of the following checks fails: a patient appears more than once; a patient appears in more than one split; a radiograph or ECG falls outside the 48-hour window; or the window extends past discharge. Imputation values, scaling parameters and class weights were estimated on the training split only. The validation split was used to select the training epoch and the decision threshold. Apart from one trial run that checked the pipeline before the main run (Section S1.15), the test split was scored only after all models had been trained; the same split was used for heart failure (after removing patients with heart failure coded earlier).

**Sample size.** No a priori sample size calculation was performed, because the study used every eligible admission in the databases. Following Riley et al. [18], we report the number of outcome events in each split and quantify the precision of every test estimate with confidence intervals.

##### S1.8 Evaluation and statistical analysis

The reported prediction of each model is the average of its five seeds' predicted probabilities. Discrimination was measured by the area under the receiver operating characteristic curve (AUROC) and the area under the precision–recall curve (AUPRC); overall accuracy of the probabilities by the Brier score; and calibration by observed versus mean predicted risk, in ten equal-width probability bins in the result files and in deciles of predicted risk (equal-count bins) in the calibration plots (Fig. 2). For threshold-based metrics, the threshold maximising Youden's J (sensitivity + specificity − 1) [19] on the validation split was applied unchanged to the test split, and sensitivity, specificity, positive and negative predictive values, F1 score and balanced accuracy are reported. Accuracy is not reported, because it is uninformative when the outcome is uncommon; AUPRC is reported because it reflects performance on the rare positive class.

Uncertainty was estimated with 1,000 bootstrap resamples of test admissions (equivalent to resampling patients), and 95% confidence intervals were taken from the 2.5th and 97.5th percentiles. Seed-to-seed variation (the mean and standard deviation of each metric over the five individual seeds) is given in the result files released with the code. Pairs of AUROCs on the same admissions were compared with DeLong's test [20] using the fast algorithm of Sun and Xu [21]. The pre-specified comparisons were: attention fusion against early fusion, late fusion, the neural EHR model and gradient boosting (all test admissions); early against late fusion; attention fusion against each of the radiograph, report and ECG models (on test admissions that had that source); and attention fusion against early and late fusion on the complete-case subset (test admissions with all four sources). Fusion models were evaluated on all test admissions and, separately, on the complete-case subset. Counts below 10 are suppressed in every reported table and figure.

**Multiplicity.** One primary hypothesis was pre-specified: attention fusion detects lung cancer better than gradient boosting on all test admissions (DeLong's test, two-sided alpha 0.05, no adjustment). All other tests were grouped into families and Holm-adjusted within each family [22], separately for each disease (all heart-failure tests are secondary): (i) the secondary model comparisons listed above, (ii) the ablations (Section S1.9), (iii) the federated comparisons (Section S1.11) and (iv) for each fairness attribute and model, the subgroup comparisons (Section S1.10). Adjusted p-values are reported in the tables (Supplementary Table S1 also gives the unadjusted values); unadjusted values for the other families and seed-to-seed standard deviations are in the result files released with the code.

##### S1.9 Ablation and sensitivity analyses

For each disease, each ablation model (Supplementary Table S8) was retrained from scratch with the same settings, and its AUROC was compared on all test admissions by DeLong's test with that of the full attention-fusion model ensembled over the same seeds (seeds 0–1, or all five where the ablation has five), so that both sides average the same number of training runs. Removing one source in turn measures what that source adds beyond the others; replacing Bio_ClinicalBERT with BERT-base tests whether clinical pretraining of the text encoder matters; and training without modality dropout tests its contribution to robustness against missing sources. The source ablations were run for both diseases and the encoder and modality-dropout ablations for lung cancer only; the ablations used the first two of the five seeds, except the lung-cancer model without the EHR source, which was available for all five (main text Table 3). In the report masking check (main text Section 3.4), 1,932 reports contained at least one of the masked word patterns.

##### S1.10 Fairness analysis

Performance was evaluated across four attributes: sex; age group (18–49, 50–64, 65–79 and 80 years or older); race and ethnicity, grouped from the free-text MIMIC-IV field into White, Black, Hispanic/Latino, Asian, other and unknown (rules in `config.RACE_RULES`); and insurance (Medicare, Medicaid, private, other, no charge, unknown) as a proxy for socioeconomic position. Race, ethnicity and insurance were never model inputs; sex and age were, as part of the EHR source. Four models were audited: attention, early and late fusion, and gradient boosting. Attention fusion is reported for both diseases and gradient boosting for lung cancer (Supplementary Tables S2, S3 and S5); results for early and late fusion, and for gradient boosting in heart failure, were similar and are available in the released result files. For every subgroup with at least 50 test admissions and 10 cases, we report AUROC, AUPRC and Brier score with bootstrap 95% CIs; calibration-in-the-large (mean predicted risk minus observed rate); and sensitivity, specificity, false-positive rate and positive predictive value at the model's single validation threshold. The threshold was deliberately not re-tuned per subgroup, so that differences in sensitivity correspond to unequal opportunity and differences in false-positive rate to unequal false alarms. Each subgroup was compared with the largest subgroup of the same attribute: the AUROC difference was tested with a z-test using DeLong variances for the two independent samples [20], Holm-adjusted within the attribute, and sensitivity and false-positive-rate differences are reported. Smaller subgroups are listed with their size but not scored. Inference in subgroups with few cases is unstable, and the smaller scored subgroups should be read descriptively.

##### S1.11 Federated learning simulation

MIMIC-IV comes from a single hospital, so five hospitals were simulated. Admissions were grouped by every combination of admission source (`admission_location`) and admission type (`admission_type`), and the admissions of each group were spread over the five sites with proportions drawn from a Dirichlet distribution with concentration 0.5 [23]. This produces sites that differ in size, outcome rate and availability of radiographs, reports and ECGs, as real hospitals do. The draw was repeated (random seed 42) until every site held at least 5% of the training admissions and at least 10 cases of the outcome in both its training and its test part (lung cancer is uncommon, so a stricter minimum could not be met). Each patient belongs to one site only. The patient-level training, validation and test splits of Section S1.7 were kept, so all methods are evaluated on the same test admissions. Site composition for lung cancer is given in Supplementary Table S4.

Four ways of training the attention-fusion model were compared:

- **Centralised:** all training data pooled (the attention-fusion model of Section S1.6). This is the reference that real hospitals cannot reach without sharing records.

- **Local only:** each site trains its own model on its own data, with early stopping on its own validation admissions; each test admission is scored by its own site's model.
- **FedAvg** [24]: in each round, every site starts from the current global weights, trains for one local epoch on its own data with a fresh AdamW optimiser, and returns its weights; the server averages them, weighting each site by its number of training admissions:

$$w^{t+1} = \sum_{k=1}^{K} \frac{n_k}{n} w_k^{t+1}$$

- **FedProx** [25]: as FedAvg, but each site minimises its local loss plus a proximal term that keeps its weights close to the global weights, which helps when sites differ:

$$\min_{w} \mathcal{L}_k(w) + \frac{\mu}{2} \|w - w^t\|^2, \qquad \mu = 0.01$$

Federated training ran for up to 50 rounds and stopped after 10 rounds without improvement. The round kept was the one with the best federated validation AUROC: each site scored its own validation admissions and the server averaged the per-site AUROCs, weighted by site size, so that no labels or predictions left a site. Platt recalibration of the federated models, by contrast, used the pooled validation labels of all sites (Section S1.15). Each method was run with three seeds (0–2); for the centralised model, the first three seeds of Section S1.6 were reused.

The methods were compared on the pooled test set and on each site's test admissions (AUROC, AUPRC and Brier score with bootstrap 95% confidence intervals, and DeLong's test for each pair of methods). The main summary is the share of the centralised gain that federated training recovers:

$$R = \frac{\text{AUROC}_{\text{federated}} - \text{AUROC}_{\text{local}}}{\text{AUROC}_{\text{centralised}} - \text{AUROC}_{\text{local}}}$$

with a bootstrap 95% confidence interval; $R$ is not interpreted when centralised and local training differ by less than 0.01 AUROC. Also reported are the number of rounds needed to reach 95% of the best validation gain over chance, and the data exchanged per round (model weights sent to and returned from every site, 32-bit floating point). The validation AUROC of every federated run, round by round, is shown in Supplementary Fig. S2. The simulation contains no differential privacy or secure aggregation, so no formal privacy guarantee is claimed; federated learning is evaluated only as a way of training without moving patient records.

##### S1.12 Explainability and its evaluation

All explanations were computed on the trained attention-fusion models and the test split; no model was retrained.

**Which sources matter.** Each test admission was re-scored with all 16 combinations of its four sources switched off, by masking their embeddings. From these scores we computed (i) the AUROC lost when each source is removed from the trained model, (ii) the AUROC obtained with each source alone, and (iii) the Shapley value [26] of each source for test AUROC:

$$\phi_j = \sum_{S \subseteq M \setminus \{j\}} \frac{|S|!\,(|M| - |S| - 1)!}{|M|!}\,[v(S \cup \{j\}) - v(S)]$$

where *M* is the set of four sources and *v*(*S*) is the test AUROC when only the sources in *S* are available. The four values add up to the AUROC with all sources minus 0.5, the AUROC of an uninformative model. The same decomposition applied to each admission's logit gives per-admission contributions, summarised as the mean absolute contribution of each source and the share of admissions in which each source contributes most. These analyses were repeated for all five seeds and are reported as mean and standard deviation.

**Which features matter within a source.** Integrated Gradients [27] was computed toward the outcome logit for the EHR features and the ECG, holding the other sources' embeddings fixed, on 1,000 randomly chosen test admissions that had an ECG (seed 0 model):

$$\mathrm{IG}_i(x) = (x_i - x_i') \times \frac{1}{m}\sum_{s=1}^{m} \frac{\partial F(x' + s - 0.5/m\,(x - x'))}{\partial x_i}$$

with $m$ = 128 steps for the EHR features and $m$ = 32 for the ECG. The EHR baseline $x'$ was the average training admission (zero for the standardised values and the training missing-rate for the indicators); the ECG baseline was a flat line. Completeness, the gap between the summed attributions and the change in logit, was checked: it was 0.0015 for the laboratory data and 0.0133 for the ECG in lung cancer, and 0.0011 and 0.0136 in heart failure. EHR attributions are summarised per feature, and ECG attributions per lead.

**Faithfulness.** Deletion curves [28] tested whether the attributions point at what the model uses. Inputs were removed in order of their attribution toward the predicted class, and the fall in the predicted-class probability was compared with removing them in random order (five random orders). For the EHR, the top 1 to 10 features were reset to the baseline; for the ECG, the signal was divided into 0.5-second segments of each lead, and 5% to 50% of the segments were set to zero in ten steps. The area over the perturbation curve (AOPC),

$$\mathrm{AOPC} = \frac{1}{K}\sum_{k=1}^{K}\left[f(x) - f(x^{(k)})\right],$$

where $f$ is the predicted-class probability and $x^{(k)}$ the input after the $k$-th deletion step, was compared between attribution order and random order, with a bootstrap 95% confidence interval for the difference. A faithful explanation gives a larger AOPC than random deletion.

**Sanity check.** Following Adebayo et al. [29], the model weights were re-initialised layer by layer from the output backwards (prediction head, fusion transformer, then the encoder of the explained source), and the attributions were recomputed on 200 of the sampled admissions after each step. Similarity to the original attributions was measured by the Spearman rank correlation of absolute attributions, averaged over admissions. An explanation that does not change when the model is randomised reflects the input rather than the model; an explanation failed the check if the correlation remained 0.5 or higher after all layers had been randomised.

**Clinician review.** The explanations were not rated by clinicians on individual patients (Section S1.15).

##### S1.13 Ethics, data access and privacy

MIMIC-IV, MIMIC-CXR, MIMIC-CXR-JPG and MIMIC-IV-ECG are de-identified databases. Their creation and sharing were approved by the Institutional Review Boards of BIDMC and the Massachusetts Institute of Technology, which waived the requirement for informed consent [3]. The analysing author completed the CITI “Data or Specimens Only Research” course, holds PhysioNet credentialed access and signed the data use agreements of the restricted databases. Separate ethics approval was not required by the University of Dhaka for this secondary analysis of de-identified, publicly available data.

In line with the data use agreements, all patient-level data stayed on the analysing author's computer. Only aggregate outputs (counts, performance metrics and summary figures) were copied to the shared project folder by a script that rejects any file containing identifier-like fields. No patient-level data, report text or images were sent to any online service, including generative AI tools. Counts below 10 are suppressed in every reported table and figure.

##### S1.14 Implementation and code availability

The pipeline was written in Python 3.11.9 with PyTorch [30] 2.14.1, scikit-learn [13] 1.9.1, SciPy [31] 1.17.1, Hugging Face Transformers [32] 5.18.0, TorchXRayVision [7] 1.5.5 and WFDB [11] (version 4.3.1 used). All experiments ran on a laptop without a graphics processor (Intel Core i7-1165G7, 16 GB RAM). The code, including automated tests on synthetic data and the leakage checks described above, will be released publicly at https://github.com/mostakphoenixsoftbd/multimodal-lungcancer-hf when the article is published and is available to editors and reviewers on request until then; the analysis plan was fixed in release v1.0-protocol, dated 5 October 2026, before the data were downloaded, and this release is archived at Zenodo (https://doi.org/10.5281/zenodo.23506613); later additions are listed in Section S1.15. The data are available from PhysioNet to credentialed users under the respective data use agreements.

##### S1.15 Deviations from the protocol

The protocol release v1.0-protocol, dated 5 October 2026, fixed the cohort definitions, the models, the hyper-parameters and the primary hypothesis. The following were added afterwards:

- **Platt recalibration** of predicted risks (Platt scaling per model and seed, fitted on the validation split). It was added after class-weighted training was seen to inflate predicted risks. Because the transformation is monotone, AUROC is unaffected; Brier scores, calibration and threshold-based metrics refer to the recalibrated risks.
- **The calibration slope summary.**
- **The tumour-word report masking analysis** (main text Section 3.4; Section S1.9).
- **Recalibration of the federated models.** One Platt model was fitted on the pooled validation split of all simulated sites. Unlike the selection of the federated round (Section S1.11), recalibration of the federated models therefore used pooled validation labels, and it is not part of the privacy-preserving protocol. AUROC is unaffected.

- **Additional analyses requested at internal review:** seed-matched ablations (Section S1.9), re-scoring in subgroups, operating points at fixed alert rates and a decision curve (Section S1.16). All of them were computed from saved predictions without retraining any model.

Before the main run, the pipeline was executed once from end to end with one seed and two training epochs, to check that it ran. This trial run also scored the test split. No setting was changed afterwards.

Two planned analyses were not carried out: the heart-failure sensitivity analysis that added NT-proBNP and troponin T to the inputs, and the clinician rating of explanations.

##### S1.16 Additional analyses

These analyses were computed from the saved predictions of the attention-fusion and gradient-boosting models; no model was retrained.

**Subgroups.** Both models were re-scored on test admissions grouped by arrival through the ED or not, by presence or absence of a chest radiograph within the window, and by the position of the outcome code: cases coded as the principal diagnosis and cases coded only as a secondary diagnosis, each compared with all test controls. The position was taken from the sequence number (`seq_num`) in the MIMIC-IV `diagnoses_icd` table and was used only to define these subsets, never as a model input. The two models were compared within each subset by DeLong's test (Supplementary Table S11).

**Operating points.** Thresholds were set on the validation split so that 5%, 10%, 20% and 30% of validation patients would be flagged, and were applied unchanged to the test split. At each threshold we report the test alert rate, sensitivity, specificity, positive and negative predictive values, and the number needed to evaluate (1/PPV, the number of flagged patients to work up per case found) (Supplementary Table S10).

**Decision curve.** The net benefit of using each model to decide who is investigated further [33] was computed on the test split over threshold probabilities of 0.5–10% for lung cancer and 5–50% for heart failure, and compared with investigating everyone (treat all) and no one (treat none, net benefit 0) (Supplementary Table S12).

Supplementary Table S6. Laboratory tests used as EHR inputs

| Test (MIMIC-IV label, blood) | Use | Missing in cohort (%) |
|---|---|---|
| Creatinine | Primary models | 0.9 |
| Urea Nitrogen | Primary models | 1.0 |
| Sodium | Primary models | 1.2 |
| Potassium | Primary models | 1.1 |
| Chloride | Primary models | 1.2 |
| Bicarbonate | Primary models | 1.3 |
| Glucose | Primary models | 1.1 |
| Hemoglobin | Primary models | 1.0 |
| Hematocrit | Primary models | 0.8 |
| White Blood Cells | Primary models | 1.0 |
| Platelet Count | Primary models | 1.0 |
| Albumin | Primary models | 53.9 |
| Magnesium | Primary models | 7.6 |
| Calcium, Total | Primary models | 10.3 |
| Anion Gap | Primary models | 1.3 |
| RDW | Primary models | 1.1 |
| NTproBNP | Extracted, not used in any model | 89.0 |
| Troponin T | Extracted, not used in any model | 81.9 |

First value within 48 h of the index time; missing = no result in that window, among all 73,637 patients.

Supplementary Table S7. Fixed settings (not tuned)

| Setting | Value |
|---|---|
| Observation window / minimum stay from index time | 48 h / 48 h |
| Split (patient-level, stratified on lung cancer) | 70% / 10% / 20%, seed 42 |

| Setting | Value |
|---|---|
| Embedding size; fusion transformer | 128; 1 layer, 4 heads, feed-forward 256, dropout 0.2 |
| Modality dropout (fusion models) | 0.2 |
| Optimiser | AdamW, learning rate 0.001, weight decay 0.0001, batch 128 |
| Epochs; early stopping | up to 30; patience 5 on validation AUROC |
| Seeds (centralised models / ablations / federated) | 5 / 2 (5 for lung cancer without EHR) / 3 |
| Gradient boosting | scikit-learn HistGradientBoosting, up to 500 iterations, balanced class weights |
| Bootstrap resamples | 1,000 (patient-level) |
| Federated sites; Dirichlet alpha; minimum site share; minimum cases per site | 5; 0.5; 5%; 10 in training and in test |
| Federated rounds; local epochs; patience; FedProx mu | up to 50; 1; 10 rounds; 0.01 |
| Integrated Gradients steps (EHR / ECG) | 128 / 32 |
| Deletion: EHR features; ECG segment; random orders | top 1–10; 0.5 s; 5 |
| Sanity-check failure threshold | Spearman rho ≥ 0.5 after full randomisation |
| Fairness: minimum subgroup size | 50 test admissions and 10 cases |

### Supplementary Tables S1–S5 and S9–S12, Supplementary Figures S1–S2

**Table S1. Pre-specified comparisons of AUROC (DeLong test).**

| Outcome | Comparison | Admissions | Family | AUROC difference | p | p (Holm) |
|---|---|---|---|---|---|---|
| Lung cancer | Attention fusion vs Early fusion | All test admissions | Secondary | -0.006 | 0.194 | 0.880 |
| Lung cancer | Attention fusion vs Late fusion | All test admissions | Secondary | +0.005 | 0.534 | 1.000 |
| Lung cancer | Early fusion vs Late | All test admissions | Secondary | +0.011 | 0.076 | 0.458 |

| | | | | | | |
|---|---|---|---|---|---|---|
| | fusion | | | | | |
| Lung cancer | Attention fusion vs Neural network (labs, age, sex) | All test admissions | Secondary | +0.079 | <0.001 | <0.001 |
| Lung cancer | Attention fusion vs Gradient boosting (labs, age, sex) | All test admissions | Primary | +0.081 | <0.001 | <0.001 |
| Lung cancer | Attention fusion vs Chest radiograph | Test admissions with radiograph | Secondary | +0.129 | <0.001 | <0.001 |
| Lung cancer | Attention fusion vs Radiology report | Test admissions with report | Secondary | +0.012 | 0.176 | 0.880 |
| Lung cancer | Attention fusion vs ECG | Test admissions with ECG | Secondary | +0.154 | <0.001 | <0.001 |
| Lung cancer | Attention fusion vs Early fusion | Complete-case test admissions | Secondary | -0.007 | 0.232 | 0.880 |
| Lung cancer | Attention fusion vs Late fusion | Complete-case test admissions | Secondary | +0.004 | 0.718 | 1.000 |
| Lung cancer | Radiology report vs Radiology report, tumour words removed (sensitivity analysis) | Test admissions with report | Report masking | +0.009 | 0.036 | 0.036 |
| Lung cancer | Attention fusion vs with tumour words removed from reports (sensitivity analysis, lung cancer) | All test admissions | Report masking | +0.015 | 0.008 | 0.016 |
| Heart failure | Attention fusion vs Early fusion | All test admissions | Secondary | -0.001 | 0.453 | 0.905 |
| Heart failure | Attention | All test | Secondary | +0.004 | 0.003 | 0.009 |

| | | | | | | |
|---|---|---|---|---|---|---|
| | fusion vs Late fusion | admissions | | | | |
| Heart failure | Early fusion vs Late fusion | All test admissions | Secondary | +0.004 | <0.001 | 0.003 |
| Heart failure | Attention fusion vs Neural network (labs, age, sex) | All test admissions | Secondary | +0.084 | <0.001 | <0.001 |
| Heart failure | Attention fusion vs Gradient boosting (labs, age, sex) | All test admissions | Secondary | +0.090 | <0.001 | <0.001 |
| Heart failure | Attention fusion vs Chest radiograph | Test admissions with radiograph | Secondary | +0.069 | <0.001 | <0.001 |
| Heart failure | Attention fusion vs Radiology report | Test admissions with report | Secondary | +0.089 | <0.001 | <0.001 |
| Heart failure | Attention fusion vs ECG | Test admissions with ECG | Secondary | +0.028 | <0.001 | <0.001 |
| Heart failure | Attention fusion vs Early fusion | Complete-case test admissions | Secondary | 0.000 | 0.892 | 0.905 |
| Heart failure | Attention fusion vs Late fusion | Complete-case test admissions | Secondary | +0.007 | 0.002 | 0.006 |

*Primary hypothesis (lung cancer: attention fusion vs gradient boosting) tested at alpha 0.05 without adjustment; all other comparisons Holm-adjusted within their family.*

#### Table S2. Performance by subgroup, lung cancer: attention fusion.

| Subgroup | Test admissions (cases) | AUROC (95% CI) | Calibration-in-the-large | Sensitivity | False-positive rate | AUROC difference vs largest group |
|---|---|---|---|---|---|---|
| Sex | | | | | | |
| Female | 7,228 (135) | 0.833 (0.798–0.864) | -0.001 | 71.1% | 20.8% | -0.001 (p = 0.957) |
| Male | 7,500 (118) | 0.834 (0.796– | 0.000 | 61.0% | 15.7% | reference |

| | | | | | | |
|---|---|---|---|---|---|---|
| | | 0.869) | | | | |
| Age group (years) | | | | | | |
| 18–49 | 2,989 (11) | 0.950 (0.905–0.982) | +0.003 | 63.6% | 5.1% | +0.163 ($p < 0.001$) |
| 50–64 | 3,931 (81) | 0.827 (0.775–0.873) | -0.002 | 71.6% | 19.8% | +0.040 ($p = 0.451$) |
| 65–79 | 4,399 (118) | 0.787 (0.739–0.832) | -0.005 | 65.2% | 23.8% | reference |
| 80+ | 3,409 (43) | 0.806 (0.737–0.874) | +0.006 | 60.5% | 20.9% | +0.020 ($p = 0.636$) |
| Race and ethnicity | | | | | | |
| Asian | 526 | not scored (too few) | | | | |
| Black | 1,767 (18) | 0.892 (0.821–0.951) | +0.003 | 61.1% | 13.4% | +0.076 ($p = 0.119$) |
| Hispanic/Latino | 619 | not scored (too few) | | | | |
| Other | 559 (15) | 0.815 (0.744–0.881) | -0.010 | 53.3% | 17.8% | -0.002 ($p = 0.969$) |
| Unknown | 993 (17) | 0.896 (0.817–0.964) | -0.001 | 76.5% | 17.3% | +0.080 ($p = 0.119$) |
| White | 10,264 (192) | 0.816 (0.782–0.848) | -0.001 | 67.2% | 19.4% | reference |
| Insurance | | | | | | |
| Medicaid | 2,087 (withheld) | 0.877 (0.825–0.924) | -0.002 | 63.6% | 14.5% | +0.088 ($p = 0.012$) |
| Medicare | 7,790 (149) | 0.789 (0.747–0.828) | +0.001 | 63.8% | 21.7% | reference |
| Other | 413 | not scored (too few) | | | | |
| Private | 4,227 (65) | 0.871 (0.828–0.910) | -0.001 | 72.3% | 14.7% | +0.082 ($p = 0.012$) |
| Unknown | withheld | not scored (too few) | | | | |

*Single validation threshold per model (not re-tuned per subgroup). Calibration-in-the-large = mean predicted risk minus observed rate. Subgroups with fewer than 50 test admissions or 10 cases are not scored; categories with fewer than 10 test admissions are not shown, and the size or case count of the smallest shown categories of that attribute is withheld so that suppressed counts cannot be derived. p-values Holm-adjusted within each attribute. Race/ethnicity and insurance are not model inputs.*

**Table S3. Performance by subgroup, lung cancer: gradient boosting.**

| Subgroup | Test admissions (cases) | AUROC (95% CI) | Calibration-in-the-large | Sensitivity | False-positive rate | AUROC difference vs largest group |
|---|---|---|---|---|---|---|
| Sex | | | | | | |
| Female | 7,228 (135) | 0.751 (0.713–0.787) | 0.000 | 65.9% | 28.8% | -0.001 (p = 0.961) |
| Male | 7,500 (118) | 0.752 (0.705–0.792) | -0.001 | 56.8% | 20.6% | reference |
| Age group (years) | | | | | | |
| 18–49 | 2,989 (11) | 0.848 (0.702–0.945) | 0.000 | 18.2% | 1.1% | +0.127 (p = 0.114) |
| 50–64 | 3,931 (81) | 0.695 (0.632–0.752) | -0.003 | 50.6% | 23.4% | -0.026 (p = 0.895) |
| 65–79 | 4,399 (118) | 0.721 (0.675–0.764) | -0.004 | 75.4% | 40.1% | reference |
| 80+ | 3,409 (43) | 0.683 (0.593–0.762) | +0.006 | 55.8% | 27.3% | -0.038 (p = 0.895) |
| Race and ethnicity | | | | | | |
| Asian | 526 | not scored (too few) | | | | |
| Black | 1,767 (18) | 0.837 (0.748–0.912) | +0.004 | 72.2% | 18.3% | +0.093 (p = 0.103) |
| Hispanic/Latino | 619 | not scored (too few) | | | | |
| Other | 559 (15) | 0.709 (0.604–0.812) | -0.011 | 46.7% | 22.1% | -0.035 (p = 0.531) |
| Unknown | 993 (17) | 0.830 (0.717–0.939) | -0.001 | 76.5% | 23.6% | +0.086 (p = 0.277) |
| White | 10,264 (192) | 0.743 (0.710–0.779) | -0.001 | 62.0% | 26.1% | reference |
| Insurance | | | | | | |
| Medicaid | 2,087 (withheld) | 0.783 (0.718–0.841) | -0.003 | 42.4% | 16.3% | +0.060 (p = 0.115) |
| Medicare | 7,790 (149) | 0.722 (0.681–0.762) | +0.001 | 67.8% | 31.7% | reference |
| Other | 413 | not scored (too few) | | | | |

| | | | | | | |
|---|---|---|---|---|---|---|
| Private | 4,227 (65) | 0.790 (0.730–0.843) | -0.002 | 60.0% | 17.6% | +0.068 (p = 0.115) |
| Unknown | withheld | not scored (too few) | | | | |

*Single validation threshold per model (not re-tuned per subgroup). Calibration-in-the-large = mean predicted risk minus observed rate. Subgroups with fewer than 50 test admissions or 10 cases are not scored; categories with fewer than 10 test admissions are not shown, and the size or case count of the smallest shown categories of that attribute is withheld so that suppressed counts cannot be derived. p-values Holm-adjusted within each attribute. Race/ethnicity and insurance are not model inputs.*

#### Table S4. Composition of the simulated hospitals (lung cancer).

| Site | Training admissions | Training cases | Test admissions | Test cases | Outcome rate | Radiograph | Report | ECG |
|---|---|---|---|---|---|---|---|---|
| 1 | 4,464 | 68 | 1,290 | 17 | 1.5% | 22.5% | 22.5% | 91.6% |
| 2 | 24,242 | 356 | 7,020 | 121 | 1.5% | 34.9% | 34.9% | 88.8% |
| 3 | 8,990 | 157 | 2,558 | 43 | 1.7% | 30.2% | 30.2% | 90.3% |
| 4 | 8,938 | 199 | 2,449 | 45 | 2.1% | 24.5% | 24.5% | 91.0% |
| 5 | 4,911 | 104 | 1,411 | 27 | 2.2% | 23.3% | 23.3% | 90.6% |

*Outcome rate over the training, validation and test admissions of each site combined; source availability as a share of the site's admissions.*

#### Table S5. Performance by subgroup, heart failure: attention fusion.

| Subgroup | Test admissions (cases) | AUROC (95% CI) | Calibration-in-the-large | Sensitivity | False-positive rate | AUROC difference vs largest group |
|---|---|---|---|---|---|---|
| Sex | | | | | | |
| Female | 7,043 (1,112) | 0.884 (0.874–0.894) | -0.004 | 84.5% | 24.2% | -0.002 (p = 0.836) |
| Male | 7,258 (1,183) | 0.886 (0.876–0.896) | +0.003 | 84.9% | 25.0% | reference |
| Age group (years) | | | | | | |
| 18–49 | 2,971 (106) | 0.911 (0.881–0.941) | +0.006 | 60.4% | 4.7% | +0.052 (p = 0.005) |
| 50–64 | 3,860 (424) | 0.885 (0.866–0.903) | +0.003 | 77.6% | 15.0% | +0.026 (p = 0.022) |
| 65–79 | 4,231 (818) | 0.859 (0.845–0.873) | -0.003 | 83.7% | 30.4% | reference |
| 80+ | 3,239 (947) | 0.812 (0.796–0.828) | -0.007 | 91.5% | 55.2% | -0.046 (p < 0.001) |

| | | | | | | |
|---|---|---|---|---|---|---|
| Race and ethnicity | | | | | | |
| Asian | 513 (49) | 0.903 (0.853–0.944) | +0.016 | 81.6% | 17.0% | +0.024 (p = 0.981) |
| Black | 1,713 (236) | 0.898 (0.879–0.917) | +0.001 | 84.8% | 20.6% | +0.020 (p = 0.286) |
| Hispanic/Latino | 606 (53) | 0.929 (0.897–0.953) | +0.015 | 83.0% | 16.1% | +0.051 (p = 0.005) |
| Other | 546 (77) | 0.894 (0.853–0.927) | +0.004 | 80.5% | 20.7% | +0.016 (p = 0.981) |
| Unknown | 984 (213) | 0.877 (0.851–0.901) | -0.011 | 87.3% | 31.6% | -0.002 (p = 0.981) |
| White | 9,939 (1,667) | 0.878 (0.870–0.887) | -0.002 | 84.7% | 25.9% | reference |
| Insurance | | | | | | |
| Medicaid | 2,052 (179) | 0.904 (0.882–0.924) | +0.009 | 78.2% | 13.8% | +0.054 (p < 0.001) |
| Medicare | 7,463 (1,713) | 0.850 (0.840–0.859) | -0.008 | 87.5% | 37.6% | reference |
| Other | 406 (40) | 0.891 (0.848–0.930) | -0.009 | 67.5% | 12.6% | +0.042 (p = 0.072) |
| Private | 4,169 (344) | 0.905 (0.886–0.920) | +0.010 | 76.4% | 12.3% | +0.055 (p < 0.001) |
| Unknown | withheld (withheld) | 0.928 (0.888–0.962) | -0.012 | 77.8% | 10.0% | +0.078 (p < 0.001) |

*Single validation threshold per model (not re-tuned per subgroup). Calibration-in-the-large = mean predicted risk minus observed rate. Subgroups with fewer than 50 test admissions or 10 cases are not scored; categories with fewer than 10 test admissions are not shown, and the size or case count of the smallest shown categories of that attribute is withheld so that suppressed counts cannot be derived. p-values Holm-adjusted within each attribute. Race/ethnicity and insurance are not model inputs.*

#### Table S9. Federated training: heart failure.

| Training | AUROC (95% CI) | AUPRC (95% CI) | Brier | AUROC range across sites | Round kept (per seed) |
|---|---|---|---|---|---|
| Centralised (pooled data) | 0.883 (0.876–0.890) | 0.634 (0.613–0.655) | 0.089 | 0.872–0.899 | – |
| Local only | 0.862 (0.855–0.869) | 0.567 (0.546–0.590) | 0.097 | 0.833–0.881 | – |
| FedAvg | 0.882 (0.875–0.890) | 0.626 (0.605–0.647) | 0.090 | 0.872–0.896 | 26, 30, 26 |
| FedProx | 0.882 (0.875–0.889) | 0.629 (0.609–0.650) | 0.089 | 0.872–0.896 | 31, 36, 37 |

*5 simulated hospitals (Dirichlet alpha 0.5); same patient-level test set for every method. Data exchanged per round: 28.0 MB. FedAvg recovered 94.7% of the centralised gain over local training (seed-averaged predictions; 95% CI 86.1% to 104.6%), and 104.4% when computed on the mean AUROC of the individual seeds. FedProx recovered 94.7% of the centralised gain over local training (seed-averaged predictions; 95% CI 86.7% to 104.4%), and 103.7% when computed on the mean AUROC of the individual seeds. FedAvg vs local only: AUROC difference +0.021, p < 0.001 (DeLong, Holm-adjusted within the federated family). FedProx vs local only: AUROC difference +0.021, p < 0.001 (DeLong, Holm-adjusted within the federated family).*

#### Table S10. Operating points at fixed alert rates (test set).

| Model | Validation alert rate | Test alert rate | Sensitivity | Specificity | PPV | NPV | Number needed to evaluate |
|---|---|---|---|---|---|---|---|
| Lung cancer | | | | | | | |
| Attention fusion | 5.0% | 5.1% | 40.3% | 95.5% | 13.7% | 98.9% | 7.3 |
| Attention fusion | 10.0% | 9.8% | 54.1% | 90.9% | 9.4% | 99.1% | 10.6 |
| Attention fusion | 20.0% | 19.0% | 66.4% | 81.9% | 6.0% | 99.3% | 16.6 |
| Attention fusion | 30.0% | 29.3% | 78.3% | 71.6% | 4.6% | 99.5% | 21.8 |
| Gradient boosting | 5.0% | 4.7% | 21.3% | 95.6% | 7.8% | 98.6% | 12.8 |
| Gradient boosting | 10.0% | 9.8% | 37.5% | 90.7% | 6.6% | 98.8% | 15.2 |
| Gradient boosting | 20.0% | 19.2% | 52.2% | 81.4% | 4.7% | 99.0% | 21.4 |
| Gradient boosting | 30.0% | 30.2% | 66.8% | 70.5% | 3.8% | 99.2% | 26.3 |
| Heart failure | | | | | | | |
| Attention fusion | 5.0% | 5.5% | 27.1% | 98.6% | 78.8% | 87.6% | 1.3 |
| Attention fusion | 10.0% | 10.9% | 47.1% | 96.0% | 69.0% | 90.5% | 1.4 |
| Attention fusion | 20.0% | 21.1% | 68.4% | 87.9% | 52.0% | 93.6% | 1.9 |
| Attention fusion | 30.0% | 30.3% | 81.0% | 79.4% | 42.9% | 95.6% | 2.3 |
| Gradient boosting | 5.0% | 5.3% | 18.8% | 97.3% | 56.9% | 86.2% | 1.8 |
| Gradient boosting | 10.0% | 10.4% | 31.1% | 93.5% | 47.9% | 87.7% | 2.1 |

| | | | | | | | |
|---|---|---|---|---|---|---|---|
| Gradient boosting | 20.0% | 20.6% | 52.2% | 85.4% | 40.7% | 90.3% | 2.5 |
| Gradient boosting | 30.0% | 30.3% | 66.0% | 76.5% | 34.9% | 92.2% | 2.9 |

*Thresholds were set on the validation split so that the stated share of validation patients is flagged, and applied unchanged to the test split. Number needed to evaluate = 1/PPV, the number of flagged patients to work up per case found. PPV, positive predictive value; NPV, negative predictive value.*

#### Table S11. Discrimination in subgroups defined by arrival route, radiograph availability and diagnosis position (test set).

| Subgroup | Test admissions | Cases | AUROC (95% CI), attention fusion | AUROC (95% CI), gradient boosting | Difference (DeLong p) |
|---|---|---|---|---|---|
| Lung cancer | | | | | |
| Arrived through the ED | 11,339 | 186 | 0.817 (0.787–0.848) | 0.722 (0.687–0.758) | +0.095 (p < 0.001) |
| Not through the ED | 3,389 | 67 | 0.878 (0.836–0.920) | 0.839 (0.796–0.882) | +0.039 (p = 0.068) |
| Without a chest radiograph | 10,212 | 157 | 0.787 (0.753–0.822) | 0.760 (0.724–0.797) | +0.027 (p = 0.087) |
| With a chest radiograph | 4,516 | 96 | 0.902 (0.870–0.934) | 0.737 (0.688–0.787) | +0.164 (p < 0.001) |
| Cases coded as principal diagnosis (vs all controls) | 14,589 | 114 | 0.864 (0.830–0.898) | 0.785 (0.745–0.826) | +0.078 (p < 0.001) |
| Cases coded only as secondary diagnosis (vs all controls) | 14,614 | 139 | 0.808 (0.773–0.844) | 0.726 (0.685–0.767) | +0.082 (p < 0.001) |
| Heart failure | | | | | |
| Arrived through the ED | 11,032 | 1,626 | 0.888 (0.880–0.897) | 0.799 (0.789–0.810) | +0.089 (p < 0.001) |
| Not through the ED | 3,269 | 669 | 0.871 (0.857–0.886) | 0.784 (0.765–0.803) | +0.087 (p < 0.001) |
| Without a chest radiograph | 9,925 | 1,572 | 0.878 (0.869–0.887) | 0.792 (0.781–0.803) | +0.086 (p < 0.001) |
| With a chest radiograph | 4,376 | 723 | 0.900 (0.889–0.912) | 0.800 (0.783–0.816) | +0.101 (p < 0.001) |
| Cases coded as principal diagnosis (vs all | 12,322 | 316 | 0.933 (0.919–0.947) | 0.828 (0.807–0.849) | +0.105 (p < 0.001) |

| | | | | | |
|---|---|---|---|---|---|
| controls) | | | | | |
| Cases coded only as secondary diagnosis (vs all controls) | 13,985 | 1,979 | 0.877 (0.870–0.885) | 0.789 (0.779–0.799) | +0.088 (p < 0.001) |

*Models re-scored from saved test predictions; no model was retrained. Principal versus secondary position of the outcome code is taken from MIMIC-IV diagnoses_icd (seq_num) and used only to define these subsets; each case subset is compared with all test controls. p-values are unadjusted; the analysis is descriptive. Lung cancer: of 253 test cases, 45.1% had the outcome coded as the principal diagnosis and 73.5% arrived through the ED (77.0% of all test admissions). Test admissions (cases) by MIMIC-IV admission type: EW EMER. 8,223 (147); OBSERVATION ADMIT 2,021 (29); SURGICAL SAME DAY ADMISSION 930 (34); URGENT 1,836 (31); other types (AMBULATORY OBSERVATION, DIRECT EMER., DIRECT OBSERVATION, ELECTIVE, EU OBSERVATION) pooled, counts not shown separately. Heart failure: of 2,295 test cases, 13.8% had the outcome coded as the principal diagnosis and 70.9% arrived through the ED (77.1% of all test admissions). Test admissions (cases) by MIMIC-IV admission type: DIRECT EMER. 524 (90); EU OBSERVATION 671 (42); EW EMER. 7,983 (1,256); OBSERVATION ADMIT 1,974 (288); SURGICAL SAME DAY ADMISSION 906 (88); URGENT 1,792 (467); other types (AMBULATORY OBSERVATION, DIRECT OBSERVATION, ELECTIVE) pooled, counts not shown separately. Counts below 10 are suppressed.*

#### Table S12. Decision-curve net benefit (test set).

| Threshold probability | Net benefit, attention fusion | Net benefit, gradient boosting | Net benefit, treat all |
|---|---|---|---|
| Lung cancer (prevalence 1.7%) | | | |
| 1.0% | 0.0105 | 0.0091 | 0.0073 |
| 2.0% | 0.0076 | 0.0056 | -0.0029 |
| 5.0% | 0.0046 | 0.0014 | -0.0345 |
| 10.0% | 0.0032 | 0.0000 | -0.0920 |
| Heart failure (prevalence 16.0%) | | | |
| 10.0% | 0.1142 | 0.0963 | 0.0672 |
| 20.0% | 0.0869 | 0.0567 | -0.0494 |
| 30.0% | 0.0677 | 0.0316 | -0.1993 |
| 40.0% | 0.0527 | 0.0161 | -0.3992 |

*Net benefit = TP/n − FP/n × pt/(1 − pt) at threshold probability pt (Vickers and Elkin), computed on the test split from saved predictions; treat none has net benefit 0. Selected thresholds shown; the full curves cover 0.5–10% (lung cancer) and 5–50% (heart failure).*

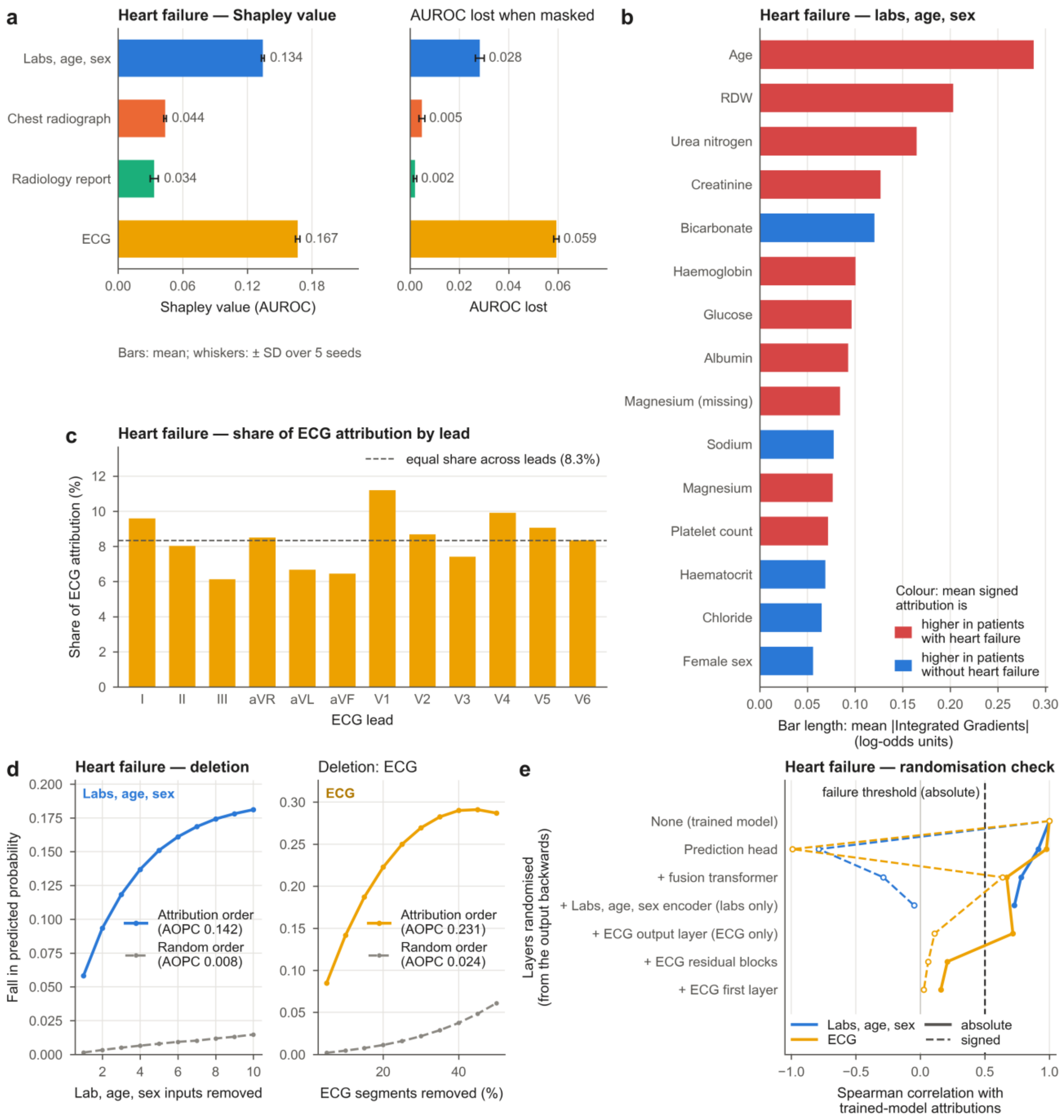


**Supplementary Fig. S1.** Explanations and their evaluation for heart failure (attention-fusion model, test set). Panels as in main text Fig. 3: (a) Shapley value of each source for AUROC and AUROC lost when the source is masked; (b) laboratory, age and sex attributions; (c) share of ECG attribution by lead; (d) deletion curves; (e) sanity check with absolute (solid) and signed (dashed) rank correlations and the failure threshold.

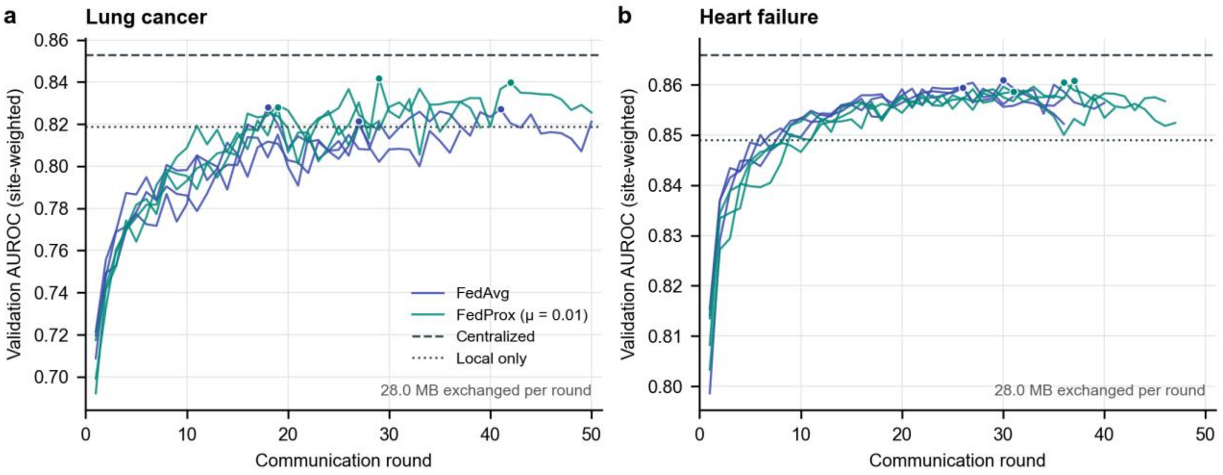


**Supplementary Fig. S2.** Federated training by round for (a) lung cancer and (b) heart failure. Site-weighted validation AUROC of the attention-fusion model after each communication round, for FedAvg and FedProx (three seeds each; dots mark the round kept). Dashed and dotted lines show centralised and local-only training. Each round exchanged 28.0 MB of model weights.
