## Supplementary material for "Explainable Multimodal AI for In-Hospital Detection of Lung Cancer and Heart Failure Using Routine Data: Development, Internal Validation, and Simulated Federated Evaluation": TRIPOD+AI Checklist

*Item wording is abridged from Collins et al., BMJ 2024;385:e078378. Section numbers refer to the main manuscript; "S" items to the Supplementary Material. D = development, E = evaluation; this study does both (internal validation).*

| Section / topic | Item | Checklist item (abridged) | Where reported |
|---|---|---|---|
| Title | 1 | Identify the study as developing or evaluating a multivariable prediction model, the target population and the outcome | Title |
| Abstract | 2 | See TRIPOD+AI for Abstracts | Abstract (structured: Objective, Methods, Results, Conclusion) |
| Background | 3a | Healthcare context (diagnostic or prognostic) and rationale | Section 1, paragraphs 1–2 |
| | 3b | Target population and intended purpose in the care pathway | Section 1; Section 3.2; Section 5.7 |
| | 3c | Known health inequalities between sociodemographic groups | Section 2.2 |
| Objectives | 4 | Study objectives, development and/or validation | Section 1, contribution list; Abstract, Objective |
| Data | 5a | Sources of data for development and evaluation | Section 3.1; Supplementary S1.2 |
| | 5b | Dates of collected data | Section 3.1 (MIMIC-IV 2008–2022; MIMIC-CXR 2011–2016; MIMIC-IV-ECG 2008–2019) |
| Participants | 6a | Key elements of the setting | Section 3.1 (BIDMC; patients admitted through the ED or to intensive care) |
| | 6b | Eligibility criteria | Section 3.2 (including exclusion of patients already known to have the disease); Fig. 1a |
| | 6c | Treatments received and how handled | Section 3.3 ("No treatment given during the window was used") |
| Data preparation | 7 | Pre-processing and quality checking, including across sociodemographic groups | Section 3.3; Supplementary S1.5; automated leakage checks (Section 3.5; Supplementary S1.7) |
| Outcome | 8a | Outcome definition, time horizon, how and when assessed | Section 3.2 (discharge ICD codes; prediction at 48 h after arrival) |
| | 8b | Qualifications of outcome assessors if subjective | Section 3.2 (coded outcome, no outcome assessor interpreted the predictors) |
| | 8c | Blinding of outcome assessment | Section 3.2 (codes assigned at discharge, independent of the model) |
| Predictors | 9a | Choice of initial predictors | Section 3.3; Supplementary S1.5 and Table S6 (routine first-48-hour data; natriuretic peptides and troponin excluded by design) |
| | 9b | Definition of all predictors, how and when measured | Section 3.3; Supplementary S1.5 |
| | 9c | Qualifications of assessors of subjective predictors | Supplementary S1.5 (radiology reports written by BIDMC radiologists; used as recorded) |
| Sample size | 10 | How the study size was arrived at, and justification | Section 3.5; Supplementary S1.7 |
| Missing data | 11 | Handling of missing data | Section 3.3 (median imputation with missingness indicators; absent sources masked, not imputed) |
| Analytical methods | 12a | How the data were used (development vs evaluation) | Section 3.5 (70/10/20 patient-level split; test split used once) |

| Section / topic | Item | Checklist item (abridged) | Where reported |
|---|---|---|---|
| | 12b | Handling of predictors (rescaling, transformation) | Section 3.3; Supplementary S1.5 |
| | 12c | Model type, rationale, building steps, hyper-parameter tuning | Section 3.4; Fig. 1b; Supplementary S1.6 and Tables S7–S8 (no tuning) |
| | 12d | Heterogeneity in performance across clusters | Section 3.6 (per-site performance in the federated simulation) |
| | 12e | Performance measures and plots, with rationale | Section 3.5; Fig. 2 |
| | 12f | Model updating (recalibration) | Section 3.4 (Platt scaling fitted on the validation split) |
| | 12g | How predictions were calculated for evaluation | Section 3.5 (average of five seeds; validation threshold) |
| Class imbalance | 13 | Class imbalance methods, why and how | Section 3.4 (class-weighted loss, then recalibration); balanced weights for gradient boosting in Supplementary Section S1.6 and Table S7 |
| Fairness | 14 | Approaches to address fairness and rationale | Section 3.5, Fairness; Supplementary S1.10 |
| Model output | 15 | Output of the model (probability, classification) | Section 3.5 (calibrated probability; binary flag at the validation threshold) |
| Training vs evaluation | 16 | Differences in setting between development and evaluation data | Section 3.5 (same centre, random patient split); Section 5.6 |
| Ethical approval | 17 | Ethics committee | Section 3.8 (BIDMC and MIT IRB approval of the databases; separate approval not required by the University of Dhaka) |
| Funding | 18a | Funding source and role | Declarations, Funding |
| Conflicts of interest | 18b | Conflicts of interest | Declarations, Declaration of competing interest |
| Protocol | 18c | Where the protocol can be accessed | Code availability (analysis plan fixed in release v1.0-protocol, dated 5 October 2026, before data download; archived at https://doi.org/10.5281/zenodo.23506613) |
| Registration | 18d | Registration | Not registered |
| Data sharing | 18e | Availability of study data | Data availability (PhysioNet, credentialed access) |
| Code sharing | 18f | Availability of analytical code | Code availability (protocol release at https://doi.org/10.5281/zenodo.23506613; complete pipeline public on publication, available to editors and reviewers on request until then) |
| Patient and public involvement | 19 | PPI in design, conduct, reporting | Section 3.8 ("There was no patient or public involvement") |
| Participants | 20a | Flow of participants, with and without outcome | Section 4.1; Fig. 1a; Table 1 |
| | 20b | Characteristics overall and per setting | Table 1; Supplementary Table S4 (per simulated site) |
| | 20c | Comparison of predictor distributions with development data | Table 1 (by split) |
| Model development | 21 | Number of participants and events in each analysis | Section 4.1; Tables 1–4; Supplementary Tables S4, S5 and S9 |
| Model specification | 22 | Details of the full model (formula, code, object) | Section 3.4; Supplementary S1.6; Code availability |
| Model performance | 23a | Performance with CIs, including key subgroups | Sections 4.2–4.3 and 4.6; Tables 2–4; Fig. 4; Supplementary Tables S1–S3, S5 and S9 |
| | 23b | Heterogeneity across clusters | Section 4.7; Table 4 (range across sites); Supplementary Table S4 |
| Model updating | 24 | Results of model updating | Sections 4.2–4.3 (calibration before and after Platt recalibration) |
| Interpretation | 25 | Overall interpretation, including fairness | Sections 5.1–5.5 |
| Limitations | 26 | Limitations | Section 5.6 |
| Usability | 27a | Handling of poor-quality or unavailable inputs in practice | Sections 3.3 and 3.4 (availability mask, modality dropout); Section 4.2 |

| Section / topic | Item | Checklist item (abridged) | Where reported |
|---|---|---|---|
| | | | (complete-case results) |
| | 27b | Whether users interact with input handling | Section 6 (inputs are taken automatically from data already in the record; no user input) |
| | 27c | Next steps for research, applicability and generalisability | Sections 5.7 and 6 |
